# *VCAM1*-expressing T cells and systemic autoimmunity in Regnase-1 deficiency

**DOI:** 10.1101/2025.01.21.25320127

**Authors:** Delphine Cuchet-Lourenço, Matilde I. Conte, Sooyeon Chang, Nika Ten, Davide Eletto, Olivier Papapietro, Vincent Plagnol, Mike de Kok, Ilie Hashim, Lourdes Ceron-Gutierrez, Marlous van den Braber, James Curtis, Harriet C. T. Groom, Mailis Maes, Rainer Doffinger, Juan Garcia Vallejo, Cecilia Dominguez Conde, Joao Farela Neves, Sergey Nejentsev

## Abstract

Autoimmunity develops as a result of a breakdown in immune tolerance and activation of autoreactive immune cells. Most of the common autoimmune diseases are polygenic^1^ suggesting dysregulation in multiple signalling pathways. By contrast, in monogenic Inborn Errors of Immunity (IEI), which also can result in autoimmunity, the disease is triggered by a single genetic defect. Therefore, the discovery of causative mutations in IEI allows tracing the molecular mechanisms leading to autoimmunity in humans from a defect in the function of a specific gene to patients’ clinical and immunological phenotype. Here, we discovered an IEI patient with systemic autoimmunity caused by a private homozygous protein-truncating mutation in gene *ZC3H12A* leading to deficiency of Regnase-1, a regulatory RNase^2–5^. Flow cytometry, bulk T cell transcriptome analysis and single-cell RNA sequencing demonstrated expansion of γδ T cells expressing VCAM-1 and IFNγ genes. We show that Regnase-1 directly targets 3’UTR of *VCAM1* and the coding sequence of *IFNG* mRNAs. These findings highlight a new autoimmunity mechanism in humans, where Regnase-1 deficiency causes expansion of *VCAM1*+*IFNG*+ T cells and their interaction with integrin α4β1-expressing B cells, which showed upregulation of IFN-response genes and activation, leading to systemic autoimmunity. Furthermore, we show that *VCAM1+* T cells are present in organs of donors and are expanded in the blood of patients with systemic lupus erythematosus, a common autoimmune disease characterised by systemic autoimmunity.

## New monogenic autoimmune disease

Patient P1 was born to a consanguineous marriage of first cousins and suffered from an autoimmune disease of unknown cause. His parents were healthy, as were his two older brothers. Soon after birth, he developed severe watery diarrhoea, splenomegaly, autoimmune anaemia and thrombocytopenia that all responded to corticosteroid treatment, and later developed autoimmune hepatitis that responded to azathioprine treatment (Fig. 1a and Supplementary Fig. 1a). He suffered from multiple recurrent respiratory infections that eventually led to bronchiectasis (Supplementary Fig. 1b) and later had meningitis caused by varicella-zoster virus (VZV). Multiple therapeutic trials were attempted, including mycophenolate, rituximab, sirolimus and tocilizumab, but the patient was refractory to these treatments and eventually developed severe myelofibrosis and transfusion dependence. When the patient received hematopoietic stem-cell transplantation (HSCT) from an HLA-matched related sibling donor after conditioning (Supplementary Fig. 1a), he had complete engraftment, and the clinical manifestations of his disease were corrected. Despite that, he developed severe skin graft versus host disease and eventually succumbed to infection.

**Figure 1.**
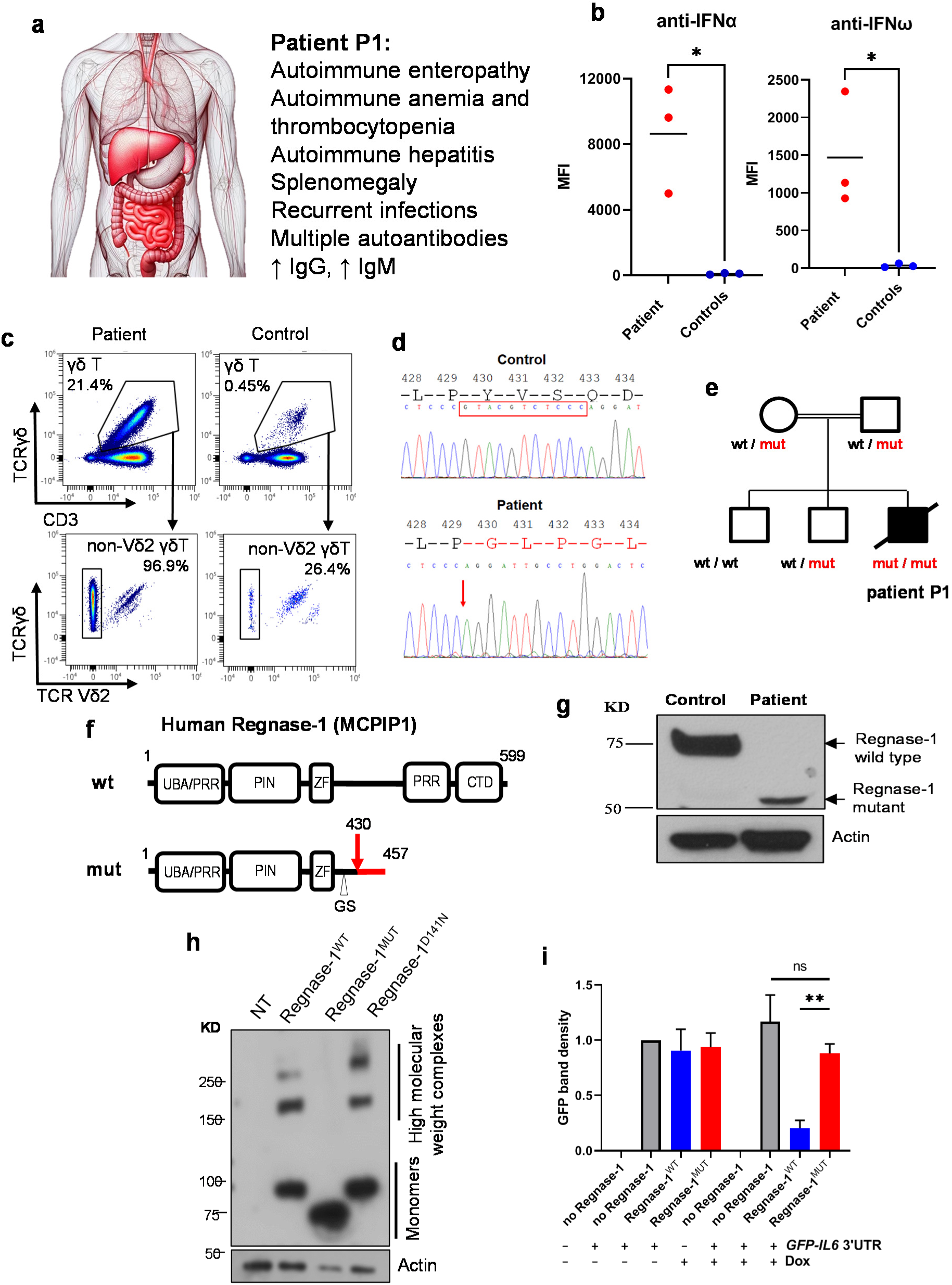
Frameshift deletion in the *ZC3H12A* gene leads to Regnase-1 deficiency and autoimmunity. (**a**) Clinical and immunological presentation of patient P1. (**b**) Anti-IFNα and anti-IFNω autoantibodies in patient P1 were measured in three different serum samples and are shown in comparison with healthy controls (N = 3). Unpaired t test, * < 0.05. MFI, Mean Fluorescence Intensity. (**c**) Flow cytometry analysis of γδ T cells in peripheral blood mononuclear cells (PBMCs). Cell frequencies are shown as % of total PBMCs (top panel) and as % of total γδ T cells (bottom panel). (**d**) Sequence chromatograms showing frameshift deletion in the *ZC3H12A* gene in patient P1. The location of the deletion is shown by red arrow; 11 deleted nucleotides are shown by red frame. (**e**) Family tree of patient P1 (wt, wild-type allele; mut, mutant allele). (**f**) Regnase-1 protein domains and motifs. Red arrow indicates frameshift mutation p.Y430Gfs*27 in patient P1. Regnase-1 is also known as Monocyte Chemotactic Protein-Induced Protein 1 (MCPIP1). (**g**) Representative immunoblot shows Regnase-1 protein in fibroblasts. N = 3. (**h**) Lysates of HEK293T cells transfected with plasmids expressing Flag-and GFP-tagged wild-type (Regnase-1^WT^), mutant (Regnase-1^MUT^) or RNase-dead (Regnase-1^D141N^) Regnase-1 proteins were studied using crosslinking with 1 mM disuccinimidyl suberate (DSS) followed by gel electrophoresis and immunoblotting with an anti-Flag antibody. Representative of 3 gels. NT, not transfected. (**i**) Regnase-1 activity assay: Regnase-1-knockout HeLa (HeLa-KO) cells either not expressing Regnase-1 or expressing exogenous Regnase-1 proteins under the control of the Dox-inducible promoter were transfected with plasmid *GFP-IL6 3’UTR*. +Dox indicates overnight incubation with 20 ng/mL doxycycline. The graph shows relative raw integrated density of the GFP band corrected for the background and actin band density ± SEM. N = 4. Unpaired two-tailed t-test with unequal variance; ** < 0.01; ns, not significant.

Throughout his life, the patient had elevated serum IgG and IgM, but absent IgA and reduced responses to both protein and polysaccharide vaccines (Supplementary Table 1). He had multiple autoantibodies, including anti-erythrocyte, anti-liver-kidney microsomal, anti-smooth muscle, anti-double stranded DNA and anti-nuclear antibodies (Supplementary Table 1). Moreover, we found that the patient had elevated anti-IFNα and anti-IFNω autoantibodies (Fig. 1b), which likely have affected his IFN-mediated antiviral responses explaining VZV meningitis and susceptibility to respiratory viral infections. Analysis of serum cytokines showed increased IL-6 and occasionally elevated IFNγ in the patient, while IL-10 and TNFα were not significantly different to controls (Supplementary Fig. 1c).

Flow cytometry analysis of peripheral blood mononuclear cells (PBMCs) showed expanded γδ T cells in the patient accounting for 21.4% of all PBMCs and 96.9% of these cells were non-Vδ2 (Fig. 1c; Supplementary Table 2). Of these cells, 54% expressed CD8 (Supplementary Fig. 2a). Furthermore, conventional CD8+ T cells were also increased in patient P1 (68.3% of T cells) and the majority of these cells had CD27–CD45RA+ cytotoxic terminally differentiated effector memory (Temra) phenotype (65.3% of CD8+ T cells; Supplementary Fig. 2b). Reciprocally, CD8+ central memory (Tcm), CD8+ naive and CD4+ naive T cells were reduced (Supplementary Table 2). We also found that the number of memory B cells was strongly reduced in the patient (Supplementary Fig. 2c), which probably have contributed to his poor responses to vaccinations (Supplementary Table 1). Similarly, numbers of mucosal-associated invariant T (MAIT) cells, natural killer cells and plasma cells were reduced in the patient, while the fraction of non-classical CD14+CD16+ monocytes was increased (Supplementary Fig. 2d-g; Supplementary Table 2).

## The new autoimmune disease is caused by Regnase-1 deficiency

Exome sequencing of patient P1 identified 158 rare variants (see Methods), of which 45 were synonymous and were unlikely to be causative. Of the remaining 113 rare variants only two were homozygous, an in-frame 6-nucleotide insertion and a frameshift 11-nucleotide deletion, both located in exon 6 of the *ZC3H12A* gene (Supplementary Fig. 3a). The 11-nucleotide deletion was the only homozygous protein-truncating variant found in the patient. No homozygous protein-truncating variants in the *ZC3H12A* gene were reported previously (e.g. such variants are absent in the gnomAD database of >800,000 subjects^6^), which suggests that they are pathogenic. The homozygous 11-nucleotide deletion co-segregated with the disease in the family (Fig. 1d,e). Taken together, these results indicated that this deletion is the disease-causative mutation in patient P1.

The human *ZC3H12A* gene encodes a broadly expressed 599 amino acid protein Regnase-1 (regulatory RNase-1) that is known to cut stem-loop RNA structures in 3’UTRs of mRNAs of several pro-inflammatory genes leading to their degradation and therefore reduced protein expression, which restrains inflammation.^2–5^ The 11-nucleotide deletion affects Regnase-1 protein at position 430 producing a new reading frame of 26 amino acids followed by a premature stop codon (p.Y430Gfs*27; Fig. 1f). In the patient’s cells, we found no full-length Regnase-1 protein, and the truncated mutant protein was present instead, but its amount was strongly reduced compared to healthy controls (Fig. 1g and Supplementary Fig. 3b). Given that *IL6* mRNA is a known Regnase-1 target^2^, we studied IL-6 secretion and found that its levels were increased in supernatants of cultured patient’s fibroblasts after LPS stimulation and after prolonged cell culture without stimulation (Supplementary Fig. 3c). Regnase-1 has been reported to form high molecular weight protein complexes via its C-terminal part^7^, which is missing in the mutant protein in patient P1. Accordingly, the mutant Regnase-1 was unable to make such complexes, contrary to the wild-type protein (Fig. 1h and Supplementary Fig. 3d,e). Furthermore, its activity against a plasmid containing *IL6* 3’UTR sequence was completely abolished (Fig. 1i and Supplementary Fig. 4). Together, these data show that the frameshift deletion in the *ZC3H12A* gene in patient P1 results in Regnase-1 deficiency.

## Dysregulated T cell transcriptome and Regnase-1 mRNA targets

Bulk RNA-seq analysis identified genes *VCAM1, SOX4*, *RHOBTB3* and *IRAK3* that were, among other genes, upregulated in unstimulated patient’s T cells as well as 4 and 24 hours after stimulation with anti-CD3/CD28. At 4 hours after stimulation, we noted increased expression of *KIT* encoding receptor tyrosine kinase c-Kit. Overexpression of *IL6* was detected 24 hours post stimulation, while the *IFNG* gene encoding IFNγ was upregulated in non-stimulated cells and 24 hours after stimulation (Fig. 2a).

**Figure 2.**
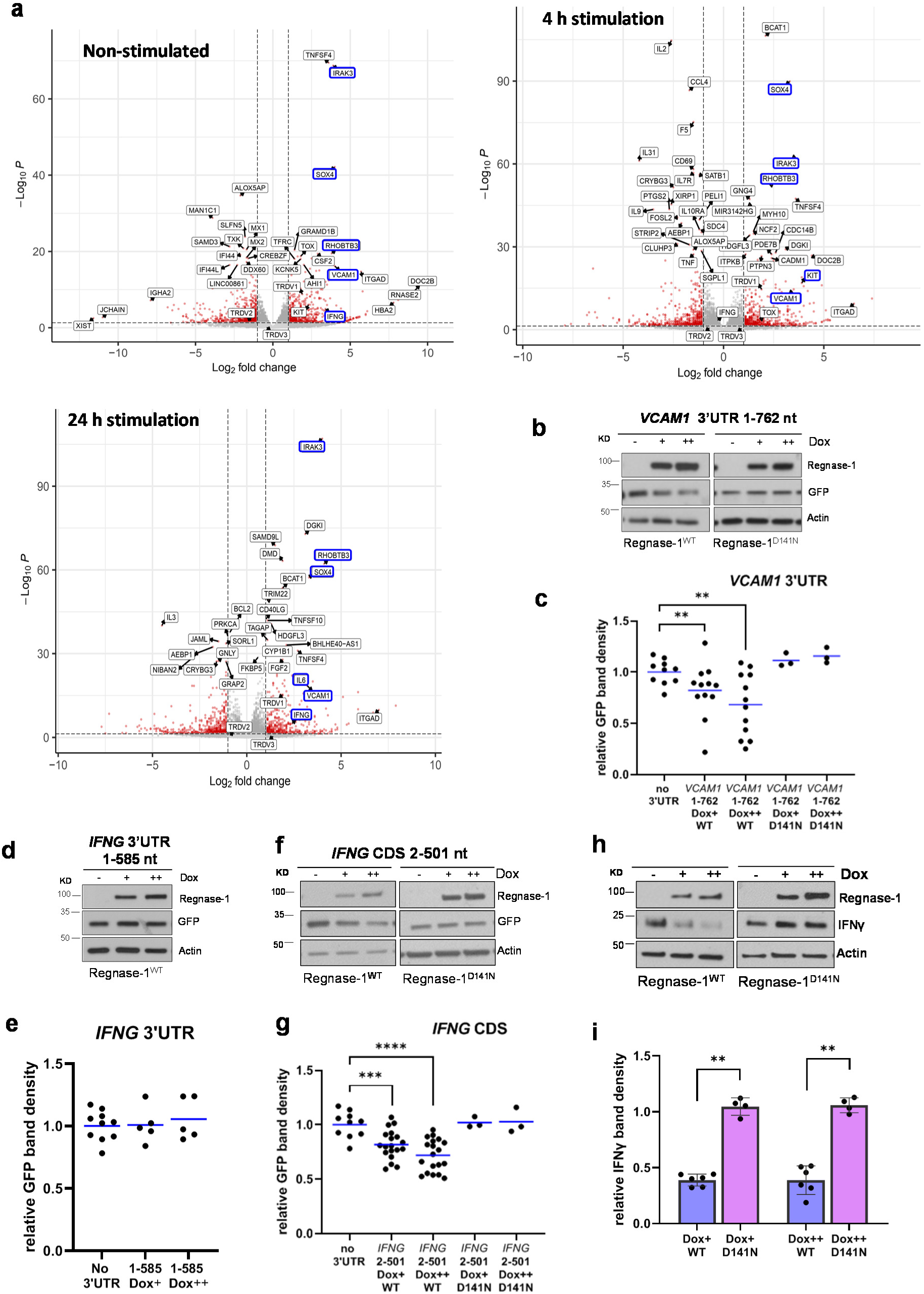
Dysregulated T-cell transcriptome and Regnase-1 mRNA targets. **(a)** Volcano plots showing differentially expressed genes in T blasts of patient P1 versus 2 healthy controls (non-stimulated, 4 and 24 hours after anti-CD3/CD28 stimulation). Genes with Log2 fold change >1 or <-1 and *P*-value <0.05 are shown as red dots. **(b, d, f)** Regnase-1 activity assay: Regnase-1-knockout HeLa (HeLa-KO) cells expressing exogenous Regnase-1^WT^ or RNase-dead Regnase-1^D141N^ under the control of the doxycycline (Dox)-inducible promoter were transfected with a plasmid encoding GFP with no 3’UTR (negative control) or GFP followed by the *VCAM1* 3’UTR 1-762 nucleotides (nt) (b), *IFNG* 3’UTR 1-585 nt (d) or by the *IFNG* CDS 2-501 nt (f). Cells were incubated overnight either without doxycycline (Dox-) or with doxycycline at final concentrations of 10 ng/mL (Dox+) and 20 ng/mL (Dox++). N ≥ 3, representative immunoblots are shown. **(c, e, g)** Band densitometry: the graphs show raw integrated density of GFP bands in Dox+ and Dox++ relative to Dox-corrected for the background and actin band density. Each dot represents a separate transfection experiment. Means are shown by blue lines. WT, Regnase-1^WT^; D141N, Regnase-1^D141N^. One-tailed Mann-Whitney test; ** < 0.01, *** < 0.001, **** < 0.0001. (**h**) HeLa-KO cells expressing Regnase-1^WT^ or Regnase-1^D141N^ under the control of the Dox-inducible promoter were transfected with a plasmid encoding IFNγ and incubated with doxycycline as above. N ≥ 4, representative immunoblots are shown. (**i**) Band densitometry: each dot represents a separate transfection experiment, bars show means. One-tailed Mann-Whitney test; ** < 0.01.

To study if transcripts upregulated in patient’s cells are direct Regnase-1 targets, we inserted 3’UTR fragments of each candidate mRNA in a plasmid downstream of green fluorescent protein (GFP) and assessed Regnase-1 activity by measuring GFP expression (Supplementary Fig. 5a-c). We found that wild-type Regnase-1 directly targets 3’UTRs of *VCAM1* (Fig. 2b,c), *SOX4,* and *RHOBTB3,* as well as *KIT* 3’UTR at nucleotide (nt) positions 1-650 (Supplementary Fig. 5d-k). This result explains the increased expression of these genes in patient’s T cells. However, we found no evidence that 3’UTRs of *IFNG* or *IRAK3* are Regnase-1 targets (Fig. 2d,e, Supplementary Fig. 6a-b).

Given that Regnase-1 recognises stem-loop RNA structures rather than specific 3’UTR sequences^2–5^, we hypothesised that it may target mRNAs also at coding sequences (CDS). Indeed, we found that Regnase-1 directly targets CDS of *IFNG* (Fig. 2f,g). Accordingly, in cells expressing wild-type Regnase-1, IFNγ protein production was lower than in cells expressing RNase-dead Regnase-1^D141N^ (Fig. 2h,i). *IRAK3* mRNA was also targeted by Regnase-1 at CDS positions 312-1791 nt (Supplementary Fig. 6c-h). This unexpected finding explained the increased expression of *IFNG* and *IRAK3* in patient’s T cells.

## scRNA-seq identifies expanded *VCAM1+* **γ**δ T and Temra cytotoxic cells

We used single cell RNA-seq (scRNA-seq) to compare PBMCs of patient P1 and 7 unrelated healthy controls, including 2 adults studied using the same platform as the patient and 5 age-matched children studied previously.^8^ Annotation by CellTypist^9^ and further curation (Supplementary Fig. 7) confirmed our flow cytometry findings, including the expansion of γδ T cells, Temra cytotoxic cells and non-classical monocytes (Fig. 3a,b; Supplementary Table 3).

**Figure 3.**
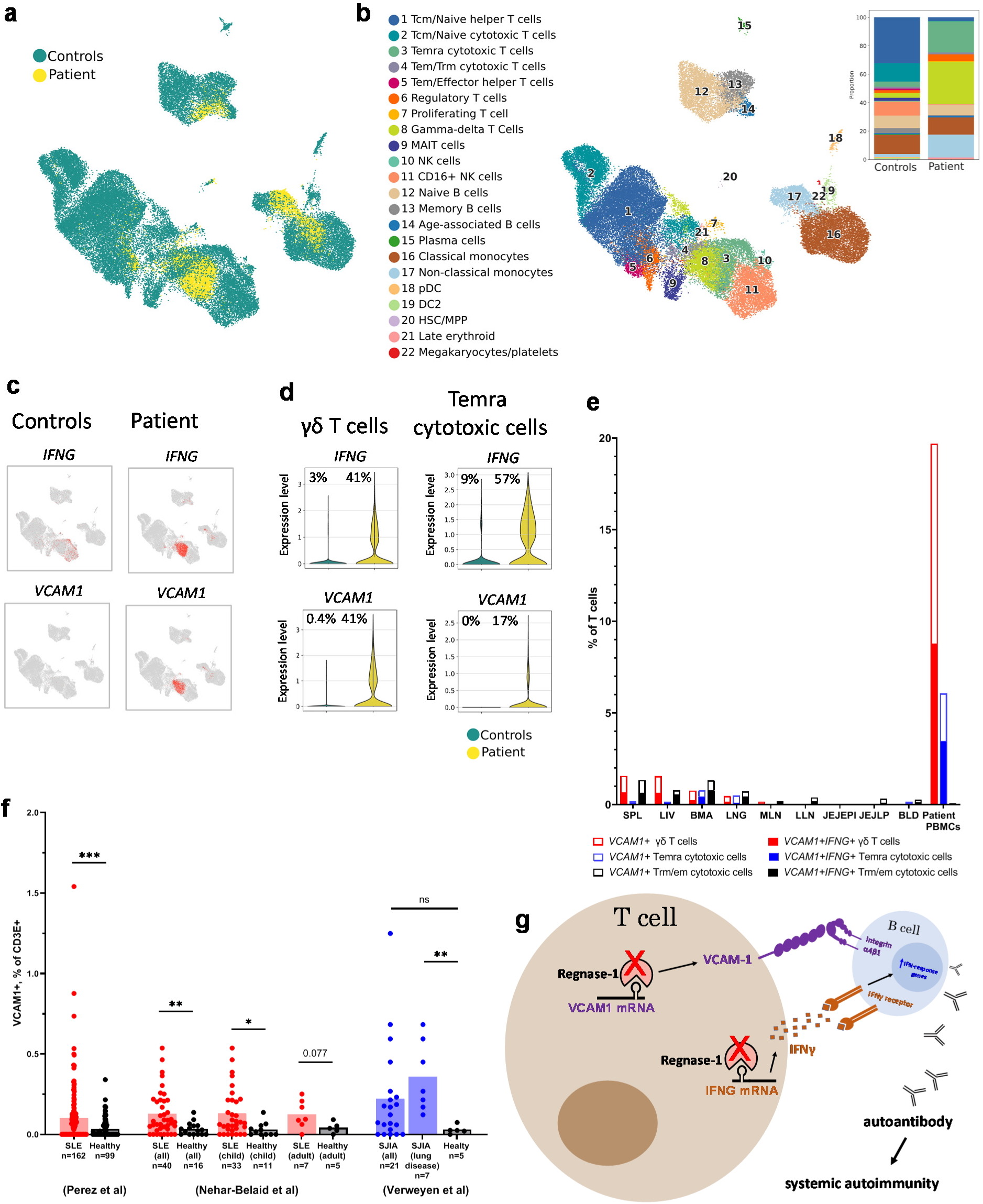
scRNA-seq identifies expanded *VCAM1+* T cells in the blood of patient P1. **(a)** Uniform Manifold Approximation and Projection (UMAP) of PBMCs of patient P1 and 7 healthy controls, including 5 age-matched controls published previously^8^ and 2 adult donors. **(b)** UMAP showing cell type annotation of PBMC clusters based on CellTypist prediction^9^ and manual curation. (Inset) The bar graph shows cell type proportions. **(c)** UMAP plots showing gene-expressing cells (expression level > 0) in red; cells where gene expression level = 0 are shown in grey. **(d)** Violin plots showing gene-expressing cells. Percentages of cells with gene expression level > 0 are shown on each plot. **(e)** *VCAM1*+ T cells in PBMCs of patient P1 and organs of the Cross-tissue Immune Cell Atlas donors. Percentages of cells with *VCAM1* expression level > 0 are shown. Data of the Cross-tissue Immune Cell Atlas donors are form https://www.tissueimmunecellatlas.org/: spleen (SPL), liver (LIV), bone marrow (BMA), lungs (LNG), mesenteric lymph nodes (MLN), lung-draining lymph nodes (LLN), blood (BLD), jejunum intraepithelial layers (JEJEPI), jejunum lamina propria (JEJLP); only organs with more than 1,000 sequenced T cells are shown. Trm/em, T resident memory/effector memory. **(f)** Frequencies of *VCAM1*+*CD3E*+ cells in the blood of patients with systemic lupus erythematosus (SLE), systemic juvenile idiopathic arthritis (SJIA) and healthy controls. Data are from scRNA-seq datasets of PBMCs.^14–16^ Percentages of cells with *VCAM1* expression level > 0 are shown. Each dot represents a subject, bars show means. Mann-Whitney test; * < 0.05, ** < 0.01, *** < 0.001. **(g)** Mechanism of systemic autoimmunity in patient P1.

Non-classical monocytes of the patient showed particularly high expression of *IRAK3*. Curiously, *IRAK3,* which normally is found in myeloid cells, was also expressed in the patient’s B and T cells, e.g. Tcm/naive and Treg cells (Supplementary Fig. 8). This explains why we detected *IRAK3* upregulation in bulk T cell transcriptomes.

On Uniform Manifold Approximation and Projection (UMAP) plots, clusters of patient’s γδ T and Temra cytotoxic cells overlapped (clusters 8 and 3, respectively; Fig. 3b), suggesting that their gene expression programmes are highly similar. *IFNG* was expressed in 41% and 57% of these cells respectively (Fig. 3c,d). Many of these cells also expressed the *ITGAD* gene (Supplementary Fig. 9), resembling *ITGAD*+ γδ T cells described previously.^9^

Notably, the *VCAM1* gene, encoding Vascular Cell Adhesion Molecule-1 (VCAM-1), was expressed by 41% of patient’s γδ T and 17% of his Temra cytotoxic cells compared to 0.4% and 0% of the control cells respectively (Fig. 3c,d). Strikingly, whilst 12.6% of all T cells in the blood of patient P1 were *VCAM1*+*IFNG*+, no single such cell was found in the blood of healthy controls (Table 1). Normally, VCAM-1 is expressed on activated endothelial cells, where it mediates leukocyte adhesion.^10^ VCAM-1 expression on synoviocytes, macrophages, follicular dendritic cells, and bone marrow stromal cells is also well recognised.^11–13^ However, the presence of a large number of *VCAM1*-expressing T cells in the patient’s blood was highly surprising.

**Table 1.** T cells expressing *VCAM1* and *IFNG*.

| **Cells in scRNA-seq** | **patient P1** | **7 controls** |  |  |
| --- | --- | --- | --- | --- |
| all T cells (clusters 1-9) | 4,994 |  | 25,531 |  |
| *VCAM1*+ | 1,314 | (26.3%) | 6 | (0.02%) |
| *IFNG*+ | 2,091 | (41.9%) | 300 | (1.2%) |
| *VCAM1*+*IFNG*+ | 630 | (12.6%) | 0 | (0.0%) |

Most of the patient’s B cells expressed activation marker *CD69* and 30% expressed *CD80* (Supplementary Fig. 10a). As expected, many B cells also expressed the *ITGA4* and *ITGB1* genes (Supplementary Fig. 10b) that encode integrin α4β1, a receptor for VCAM-1, which would facilitate interactions between such B cells and T cells of the patient that express *VCAM1*. Similarly, genes *IFNGR1* and *IFNGR2* encoding IFNγ receptor were expressed in B cells (Supplementary Fig. 10c), suggesting that these cells are responsive to IFNγ produced by γδ T and Temra cytotoxic cells in the patient. Accordingly, more B cells in the patient than in controls showed expression of the IFN-response genes *EIF2AK2, EPSTI1, STAT1* and *MX1* (Supplementary Fig. 10d), and this expression was associated with the expression of the integrin α4β1 genes in the same cells (Supplementary Fig. 10e).

## *VCAM1*-expressing T cells in organs of deceased donors and in the blood of patients with common autoimmune diseases

While *VCAM1*-expressing T cells appear to be extremely rare in the blood of healthy subjects, such cells may potentially exist in other organs. We analysed data of deceased donors in the Cross-tissue Immune Cell Atlas and found that *VCAM1*+ T cells, including γδ T and Temra cytotoxic cells, were present in the spleen, liver, bone marrow, and other organs at low frequencies (Fig. 3e). Given the autoimmunity phenotype of patient P1, we also studied if *VCAM1*-expressing T cells are expanded in the blood of patients with common autoimmune diseases. We analysed three published scRNA-seq datasets comprising adult and child patients with systemic lupus erythematosus (SLE) and systemic juvenile idiopathic arthritis (SJIA).^14–16^ We found that some of the patients had clearly increased frequency of *VCAM1*+*CD3E*+ cells and, overall, such cells were significantly enriched among adult and child SLE patients and among SJIA patients with lung disease (Fig. 3f and Supplementary Table 4).

## Discussion

Regnase-1 has been extensively studied in mouse models^2–5,17–19^, but the human disease caused by Regnase-1 deficiency remained unknown. Here, we report a patient with a homozygous frameshift deletion in the *ZC3H12A* gene, which, for the first time, revealed the clinical and immunological phenotype of Regnase-1 deficiency in humans. Our patient presented with systemic autoimmunity, which resembles the severe autoimmunity phenotype of *Zc3h12a*-knockout mice. Immunologically, both human and mouse Regnase-1 deficiencies manifested with hypergammaglobulinemia and multiple autoantibodies owing to B cell activation.^2,4,17^

Regnase-1 deficiency resulted in dysregulated expression of multiple mRNAs in the patient’s T cells. We show that Regnase-1 targets mRNAs not only at 3’UTRs but also at coding sequences. This finding will expand the range of genes regulated by Regnase-1. Thus, increased IFNγ expression has been consistently found in T cells of the Regnase-1-knockout mice^2,20^, but the reason for this remained unclear, given that 3’UTR of the IFNγ mRNA is not a Regnase-1 target. Our finding that instead Regnase-1 targets the coding sequence of its mRNA uncovers a new mechanism of post-transcriptional regulation of this important cytokine.

γδ T cells were expanded in the blood of the Regnase-1-deficient patient. γδ T cells have been strongly implicated in autoimmunity, as they can interact with and directly provide non-cognate help to B cells independently of other cell types leading to autoantibody secretion.^21–23^ Owing to Regnase-1 deficiency, γδ T cells of the patient frequently expressed the *IFNG* gene, and it is known that IFNγ can activate B cells promoting autoantibody production and systemic autoimmunity.^24^ Importantly, in our patient, many of the γδ T cells and transcriptionally similar Temra cytotoxic cells were unusual, expressing the adhesion molecule VCAM-1 gene. This finding was unexpected and suggested that such cells would interact with B cells that express integrin α4β1, a VCAM-1 receptor. Taken together, this highlights a pathogenic mechanism of autoimmunity in the Regnase-1-deficient patient (Fig. 3g), where VCAM-1 expressed on T cells would facilitate their interaction with B cells, bringing these cells in close proximity and making stimulation of B cells by IFNγ particularly effective. This would result in B-cell activation, autoantibody production and systemic autoimmunity. Considering that other leukocytes also express integrin α4β1, their interactions with *VCAM1*-expressing T cells may further contribute to the pathogenesis.

Our finding suggests a new mechanism of autoimmunity in humans. Given that VCAM-1 is upregulated by NF-κB signalling^10,25^, keeping VCAM-1 expression in T cells under control during inflammation must be critical for preventing systemic autoimmunity and our data show that Regnase-1 acts as an important safeguard regulating VCAM-1 expression in T cells. Interestingly, we found that *VCAM1*-expressing T cells are not exclusive to the Regnase-1-deficient patient and normally are present at low numbers in spleen, liver, bone marrow and other organs, where they may be kept away from interacting with B cells. However, in some SLE patients, *VCAM1*-expressing T cells are expanded in the blood, where such cells may freely interact with other cells expressing integrin α4β1. This may lead to B cell activation, similarly to the mechanism in the Regnase-1-deficient patient, and production of autoantibodies, a hallmark of SLE. Given these similarities, future studies should investigate the role of *VCAM1*-expressing T cells in the development of SLE and other common autoimmune diseases, which may open way to anti-VCAM-1 therapies.

## Data Availability

All data produced in the present study are available upon reasonable request to the authors

## Acknowledgements

We thank Georgina Okecha for her help with cell lines. S.N. was supported by the Wellcome Trust Senior Fellowship (095198/Z/10/Z), MRC Programme grant (MR/M012328/1), ERC Starting grant (260477) and ERC Advanced grant (832721). S.N. and R.D. are supported by the National Institute for Health Research (NIHR) Cambridge Biomedical Research Centre.

## Author contributions

D.C.-L., S.C., N.T., D.E., O.P., I.H., H.C.T.G. and M.M. performed cell experiments and analyzed the data. V.P. analyzed exome data. J.C. performed sequencing. M.C., C.D.C. and M.d.K. analyzed RNA-seq data. M.v.d.B. and J.G.V. performed flow cytometry analysis and analyzed the data. J.F.N. looked after the patient and collected the patient’s data. R.D. and L.C.-G. performed analyses of cytokines and autoantibodies. S.N. planned the experiments and analyzed the data. D.C.-L. and S.N. wrote the first draft of the manuscript.

## Methods

### Ethics

The nature and possible consequences of the study were explained to the participants and all material was obtained with informed consent in accordance with the Declaration of Helsinki and with approval from the Research Ethics committee (15/WS/0019).

### Exome sequencing and bioinformatics analysis

We isolated DNA samples from blood or peripheral blood mononuclear cells (PBMCs). Library preparation, exome capture and sequencing were done according to the manufacturers’ instructions. For exome target enrichment, Agilent SureSelect 50 Mb kit was used. Sequencing was done using Illumina HiSeq 2000. FASTQ files were aligned to the hg19 human genome reference sequence using Novoalign version 2.07.19, including hard and soft clipping, quality calibration and adapter trimming. Duplicate reads were excluded using the PICARD tool MarkDuplicates. Calling was performed using SAMtools v0.18 and single sample calling. The resulting calls were annotated with the software ANNOVAR. Candidate variants were filtered based on function: loss-of-function, non-synonymous or potential splicing altering variants (defined as being with 5 bp of the actual splice site) and frequency. To identify rare variants, we excluded known polymorphisms previously detected in 1000 Genomes^26^ and 6,500 exomes^27^. To identify large deletions and duplications we used software ExomeDepth^28^ (http://cran.r-project.org/web/packages/ExomeDepth/index.html).

### *ZC3H12A* gene mutation sequencing

To study the *ZC3H12A* gene by Sanger sequencing we used primers 5’TCATCCCAGTCCAGCTCTCT3’ and 5’TGGGAGCTCAGATCCATAGG3’ to amplify part of exon 6 and then sequenced the amplicon using one of the primers.

### Measuring anti-cytokines binding antibodies

Recombinant human cytokines were covalently coupled to different magnetic beads regions (Luminex, Netherlands). Beads were first activated with 1-ethyl-3-[3-dimethylaminopropyl] carbodiimide hydrochloride (Pierce/Thermo Fisher Scientific, #11851335) in the presence of N-hydroxysuccinimide (Pierce/Thermo Fisher Scientific, #10391314), according to the manufacturer’s instructions. The activated beads were then incubated with their correspondent cytokine at a concentration of 20 µg/ml in the reaction mixture for 2 hours at room temperature on a rotator. Beads were washed and re-suspended in blocking buffer (10 mM PBS, 1% BSA, 0.05% NaN3). Successful coupling of the cytokines to their bead set were verified with specific monoclonal antibodies.

Cytokines-coupled beads were incubated with patient sera for 1 hour in 96-well flat-bottom plate (Greiner Bio One #655096) at room temperature in the dark on an orbital shaker. Plate was placed on a magnet and buffer was aspirated. Beads were washed three times with 10 mM PBS/0.05% Tween 20. Beads were then incubated for 30 min with PE-labeled anti–human IgG-Fc antibody (Leinco/Biotrend; # I-127), washed as described, and re-suspended in 100 µl PBS/Tween. They were then analysed on a Luminex 200 analyser according to the manufacturer’s recommendations, using Bio-Plex Manager 6.1 software for analysis.

### Measuring cytokines levels

We measured levels of cytokines IL-6, IFNγ, IL-10 and TNFα in serum and levels of IL-6 in fibroblast supernatants. Serum was diluted 1:2 in RPMI. We used particle-based flow cytometry with a Human Magnetic Luminex Assay kit (R&D systems # LXSAHM) according to the manufacturer’s instructions. Briefly, standards and sample were added to a 96-well plate, following by the microspheres; plate was then incubated for 2 hours at room temperature on an orbital microplate shaker set at 700 rpm. After the incubation, the beads were washed three times with wash buffer and plate was incubated with biotinylated anti-cytokine antibodies for 60 minutes at room temperature on the orbital microplate shaker as before. Beads were washed and 50 µ L of Streptavidin-PE (SA-PE) were added to each well; plate was incubated for 30 minutes at room temperature on orbital microplate shaker as before. The beads were then washed and re-suspended in wash buffer. Cytokine levels were measured with a Luminex 200 analyser (Bio-Plex, Bio-Rad Laboratories), according to the manufacturer’s recommendations, using Bio-Plex Manager 6.1 software for analysis.

### Cells

Human primary dermal fibroblasts of patient P1 and a healthy adult control (HDFa, C0135C, ThermoFisher Scientific, UK) as well as human embryonic kidney HEK293T cells and human epithelial HeLa cells were cultured in DMEM (61965-026, Gibco, Life Technologies, UK) supplemented with 10% heat-inactivated foetal bovine serum (FCS-SA/500, Labtech), 20 µM HEPES buffer (H0887, Sigma, UK), and 100 U/mL penicillin and 100 µg/mL streptomycin (P0781, Sigma, UK).

Human PBMCs were isolated from fresh peripheral blood by centrifugation with a gradient of Ficoll*-*paque Plus (17-1440-03, GE Healthcare, UK) at 2000 rpm for 23 minutes at room temperature. Buffy coat layer containing PBMCs was washed with PBS 3 times. PBMCs were stored in liquid nitrogen.

### Cloning

The plasmid Flag-Regnase-1-pcDNA3.1 was kindly provided by the laboratory of Dr S. Akira. Direct mutagenesis was used to generate Regnase-1^MUT^ and Regnase-1^D141N^. To generate Regnase-1^D141N^, a one-step procedure was used with primers F1 and R1.

F1: 5’-AGACCAGTGGTCATCAATGGGAGCAACGTGG-3’

R1: 5’-CCACGTTGCTCCCATTGATGACCACTGGTCT-3’

A three-step procedure was used to generate Regnase-1^MUT^, corresponding to mutant Regnase-1 found in the patient. In Step 1, the 11-nucleotides deletion mutation was generated using primer F2 and R2. Then, the 6-nucleotides insertion mutation was completed in Step 2 using primers F3 and R3 and in Step 3 using primers F4 and R4.

F2: 5’-GCTGGACTCACTCCCAGGATTGCCTGGACT-3’

R2: 5’-AGTCCAGGCAATCCTGGGAGTGAGTCCAGC-3’

F3: 5’-GGGCAGTGGCAGTGGCAGCTTTGGGCC-3’

R3: 5’-GGCCCAAAGCTGCCACTGCCACTGCCC-3’

F4: 5’-GCAGTGGCAGTGGCAGCAGCTTTGGGCCC-3’

R4: 5’-GGGCCCAAAGCTGCTGCCACTGCCACTGC-3’

We sequenced the plasmids, and Sanger sequencing confirmed successful mutagenesis. We also confirmed by CLUSTALW alignment that the Regnase-1^MUT^ sequence in the pcDNA3.1 plasmid matched to the sequence read generated from exon 6 of the *ZC3H12A* gene in DNA isolated from the patient’s cells.

A two-step approach was then used to clone the superfolder GFP^29^ at the C-terminus of the Flag-Regnase-1^WT^, Flag-Regnase-1^MUT^ and Flag-Regnase-1^D141N^. During the first step, the canonical STOP codon (TAA) of each protein was changed into GAA (encodes amino acid Glu). For the Regnase-1^WT^-expressing and Regnase-1^D141N^-expressing plasmids, we used primers F5 and R5. For the Regnase-1^MUT^-expressing plasmid, we used primers F6 and R6. F5: 5’-GGGCCCTCTAGATTCCTCACTGGGGTGCT-3’

R5: 5’-AGCACCCCAGTGAGGAATCTAGAGGGCCC-3’

F6: 5’-AAGGGCGAATTCGGGAGTGG-3’

R6: 5’-GCTCTAGATTCCCAGGGCCTCCTCCTCGAACC-3’

The plasmid encoding superfolder GFP was kindly provided by Dr Erik Snapp of the Albert Einstein College in New York, USA. The superfolder GFP was flanked with XbaI sites by PCR using primers F7 and R7.

F7: 5’-GCTCTAGAGTGAGCAAGGGCGAGGAGCT-3’

R7: 5’-GCTCTAGATTACTTGTACAGCTCGTCCAT-3’

Superfolder GFP was then cloned in the unique XbaI site generating plasmids pcDNA3.1-Flag-Regnase-1^WT^-GFP, pcDNA3.1-Flag-Regnase-1^D141N^-GFP and pcDNA3.1-Flag-Regnase-1^MUT^-GFP. The Flag-and GFP-tagged Regnase-1 constructs were then cloned into the lentiviral vector pLVX-TRE3G (Clontech/Takara) via the unique sites BamHI and EcoRI, generating pLVX-TRE3G-Flag-Regnase-1^WT^-GFP, pLVX-TRE3G-Flag-Regnase-1^D141N^-GFP and pLVX-TRE3G-Flag-Regnase-1^MUT^-GFP. Compatible restriction sites for that were generated by PCR using primers F8 and R8.

F8: 5’-AAGCTTGGTACCGAGCTCGGATCCACC-3’

R8: 5’-CCGCAATTGTTACTTGTACAGCTCGTCCATGC-3’

All modified parts on each plasmid were validated by Sanger sequencing.

A two steps approach was used to clone 3’UTR sequences of the *VCAM1*, *IFNG, IRAK3, KIT, RHOBTB3,* and *SOX4* mRNAs instead of the 3’UTR sequence of the *IL6* mRNA, originally present in the plasmid *GFP-IL6 3’UTR*^30^. During the first step, the plasmid *GFP-IL6* 3’UTR was digested with EcoRI and XhoI removing the *GFP-IL6* 3’UTR sequence. Then, GFP coding sequence was amplified and flanked with the existing EcoRI site on the N-terminal part and a new added XhoI site on the C-terminal part using primers F10 and R14.

F10: 5’-GTGTGGTGGAATTCGCCACC-3’

R14: 5’-TGACGATCTCGAGCTACACATTGATCCTAACAGAAGCACA-3’

*GFP* was then reintroduced in the previously digested plasmid, generating plasmid GFP-XhoI. Then, fragments of 3’UTRs of the *VCAM1*, *IFNG, IRAK3, KIT, RHOBTB3,* and *SOX4* mRNAs were amplified by PCR using human genomic DNA and, respectively, primers F11 to F27 and R15 to R31.

F11 (VCAM1): 5’-TACTAGACTCGAGCTAATGCTTGATATGTTCAACTGGAGA-3’

R15 (VCAM1): 5’-TACAATCGGGCCCTTCTAATTCCAGAAATTATCACTTTAC-3’

F16 (IFNG): 5’-TACTAGACTCGAGTGAATGTCCAACGCAAAGCA-3’

R20 (IFNG): 5’-TACAATCGGGCCCAGTCCAACACAGCATAAGTAGT-3’

F17 (IRAK3 part 1): 5’-ACGTTCACTCGAGAGAGCAGCTGTTCCTCCAAA-3’

R21 (IRAK3 part 1): 5’-TCCACGTCCCAATTCCAGAA-3’

F18 (IRAK3 part 2): 5’-ACGTTCACTCGAGATTTCACTGGACCCACTCAT-3’

R22 (IRAK3 part 2): 5’-TACAATCGGGCCCAGGTCACTGCCAGCTTATCT-3’

F19 (IRAK3 part 3): 5’-ACGTTCACTCGAGGATGTTGGTTAGCACAGAGT-3’

R23 (IRAK3 part 3): 5’-TACAATCGGGCCCTTCCATCTGCACTGTAGCCC-3’

F20 (IRAK3 part 4): 5’-ACGTTCACTCGAGCCATCCAAACTTCCATTCATTGA-3’

R24 (IRAK3 part 4): 5’-TACAATCGGGCCCGTTTCAAACACACATGAGTCTGA-3’

F21 (KIT part 1): 5’-TACTAGACTCGAGGCAGAATCAGTGTTTGGGTCA-3’

R25 (KIT part 1): 5’-TACAATCGGGCCCGCCTTCTCTCCAATTCAATGCT-3’

F22 (KIT part 2): 5’-TACTAGACTCGAGAGCATTGAATTGGAGAGAAGGC-3’

R26 (KIT part 2): 5’-TACAATCGGGCCCATGTGAAAAGAAACAAGAGCAGT-3’

F23 (KIT part 3): 5’-TACTAGACTCGAGAGTACCTGAAAAGTAACTTGGCT-3’

R27 (KIT part 3): 5’-TACAATCGGGCCCTCTCAACAATCAAATGACTCCAC-3’

F25 (RHOBTB3) 5’-TACTAGACTCGAGCGTTGCTTAGTAATGTAACCTGG-3’

R29 (RHOBTB3) 5’-TACAATCGGGCCCTGCTGCTACACAAGTTACACG-3’

F26 (SOX4) 5’-TACTAGACTCGAGCTCGAGTCCAGCATCTCCAA-3’

R30 (SOX4) 5’-TACAATCGGGCCCACTGCCCCTGTACCACATC-3’

In addition, fragments of CDS of *IFNG* and *IRAK3* were amplified by PCR using DNA gblocks and common primers F28 and R32.

F28 5’-TACTAGACTCGAGTCTTGGTTCTCAGCTGGTTCT-3’

R32 5’-TACAATCGGGCCCTGCTCAAGTGATTCAAGGTTGT-3’

PCR products were then digested using XhoI and ApaI. Each digested PCR product was then inserted in between unique XhoI and ApaI sites present in the plasmid GFP-XhoI, generating plasmids where *GFP* is followed by sequences covering 3’UTRs of *VCAM1* nucleotides 1-762, *IFNG* nucleotides 1-585, *IRAK3* nucleotides 1-1656 (part 1), *IRAK3* nucleotides 1644-3027 (part 2), *IRAK3* nucleotides 2896-4463 (part 3), *IRAK3* nucleotides 4370-6391 (part 4), *KIT* nucleotides 1-650 (part 1), *KIT* nucleotides 629-1236 (part 2), *KIT* nucleotides 1179-2158 (part 3), *RHOBTB3* nucleotides 1-667, *SOX4* nucleotides 1-970, as well as *INFG* CDS nucleotides 2-501, *IRAK3* CDS nucleotides 2-366 (part 1), *IRAK3* CDS nucleotides 312-902 (part 2) or *IRAK3* CDS nucleotides 854-1791 (part 3).

The coding sequence of IFNγ preceded by the Flag sequence was amplified by PCR using DNA gblocks and primers F29 and R33.

F29 5’-TCTTGGTTCTCAGCTGGTTCT-3’

R33 5’-TGCTCAAGTGATTCAAGGTTGT-3’

The PCR product was digested using EcoRI and ApaI and then inserted in between unique EcoRI and ApaI sites in the plasmid *GFP-IL6 3’UTR*, thereby removing the *GFP-IL6* 3’UTR and introducing instead the following sequence: ATG -*Flag* CDS - *IFNG* CDS nucleotides 4-501.

All modified parts on each plasmid were validated by Sanger sequencing.

### Generation of the Regnase-1-knockout HeLa (HeLa-KO) cells

The Alt-R CRISPR-Cas9 system from IDT was used as recommended. CRISPR RNA (crRNA) used were: Alt-R CRISPR Negative Control crRNA (#1072544) and a human Regnase-1 specific crRNA:

/AlTR1/rCrGrGrCrCrCrGrArCrGrUrGrCrCrCrArUrCrArCrGrUrUrUrUrArGrArGr CrUrArUrGrCrU/AlTR2/

crRNA:tracrRNA (#1072532) complexes were complexed with the Cas9 nuclease (Alt-R S.p. Cas9 Nuclease V3, #1081058) before being delivered into HeLa cells. After 2 days, cells were plated as single clones. Efficient knockout of the *ZC3H12A* gene in each isolated single-cell clone was confirmed by the analysis of Regnase-1 protein expression using western blotting. HeLa clones that had Regnase-1 knocked out (HeLa-KO cells) were expanded and cryopreserved.

### Transduction of HeLa-KO cells

Lentivirus stocks were prepared by transient transfection of 3 million HEK293T cells with 13.5 µL of the transfection reagent Lipofectamine 2000 (#11668019, ThermoFisher) along with the envelope plasmid pCMV-VSV-G (1 µg, AddGene #8454), the packing plasmid psPAX2 (1.4 µg, AddGene # 12260), the expression plasmid pLVX-EF1a-Tet3G (expressing the Tet-On 3G transactivator; 2 µg) and 2 µg of plasmid pLVX-TRE3G-GFP or plasmid pLVX-TRE3G-Flag-Regnase-1-GFP expressing either Regnase-1^WT^ or one of the Regnase-1 mutants. The following morning, the media was replaced. The medium was harvested at 48 hours post transfection, cleared by low-speed centrifugation (1200 rpm, 5 min), and filtered through 0.45 µm pore size filters. Fresh collected virus stocks were used for transduction experiments (transactivator + constructs). Transductions of the HeLa-KO cells were carried out by infection in the presence of 8 µg/ml of Polybrene (107689, Sigma). After 48 hours, the virus-containing media was replaced by fresh media containing 1 µg/mL of puromycin. Two days later, the selected positive transduced cells were amplified and then selected using fresh media containing 700 µg/mL G418. Two weeks later the selected positive double-transduced cells were expanded.

### Analysis of Regnase-1 high-molecular weight complexes

To study Regnase-1 oligomerisation, the HEK293T cells were plated at 200,000 cells/well in 24-well plates and were transfected the next day using Lipofectamine 2000 (Invitrogen) with 5 µg of plasmids expressing pcDNA3.1-Flag-Regnase-1^WT^-GFP, pcDNA3.1-Flag-Regnase-1^D141N^-GFP or pcDNA3.1-Flag-Regnase-1^MUT^-GFP. The transfected HEK293T cells were cultured for 24 hours. To study Regnase-1 oligomerisation, HeLa-KO cells expressing doxycycline (Dox)-inducible Regnase-1 proteins were plated at 200,000 cells/well in 24-well plates in presence of 20 ng/mL doxycycline and cultured for 24 hours.

For studying on a native gel, the cells were lysed at 4°C for 30 min in Mammalian Protein extraction reagent (M-PER, Thermo Scientific, 78501) supplemented with proteasome inhibitor. Samples were then collected and centrifuged at 15000 g for 10 min at 4°C. The supernatants were mixed v/v with sample buffer (Bio-Rad 161-0738) before been loaded on gel (4-15% Mini-Protean TGX stain-free protein gel, Bio-Rad 4568084). The migration at 50 Volts was done in buffer (10x Tris/Glycine, cold) for 5 hours.

For crosslinking, the cells were washed and incubated at room temperature for 30 min with 1 mM disuccinimidyl suberate (DSS, CAYMAN Chemical Company, 13008). The cells were then quenched in 100 mM Tris buffer solution for 15 min at room temperature and then the proteins were extracted. The cells were lysed in lysis buffer (PIERCE, 87788) in presence proteasome inhibitor (cOmplete Mini, Roche, 11836153001) on ice for 30 min before been centrifuged at 1300 rpm at 4°C. The supernatants were then mixed v/v with x2 Laemmli sample buffer (BIO-RAD, 161-0737) containing 50 mM DTT (Merk, 1019777700). Samples were heat boiled before being loaded on 4-15% Acrylamide/bis gels (4568085, Bio-Rad).

### Plasmid transfection

To study Regnase-1 activity, HeLa-KO cells expressing doxycycline-inducible Regnase-1 proteins were plated at 1.3 x 10^5^ cells per well in a 24-well plate and transfected the next day using Lipofectamine 3000 (Invitrogen) with 1.6 µg of plasmid *GFP-IL6 3’UTR* or plasmid *GFP-IL6 5’UTR* ^30^, or one of the plasmids where GFP coding sequence was followed by a 3’UTR of the *VCAM1*, *IFNG, IRAK3*, *KIT, RHOBTB3* or *SOX4* mRNA or a CDS of *IFNG* or *IRAK3* mRNA, or a plasmid encoding IFNγ. The cells were incubated overnight either without doxycycline (Dox-) or with the final doxycycline concentrations of 10 ng/mL (Dox+) or 20 ng/mL (Dox++). On the following day, the cells were washed and lysed and proteins were extracted.

### Western blotting

Cells were washed with PBS x1 and lysed with ice-cold RIPA lysis buffer x2 (R0278, Sigma) supplemented with proteasome inhibitor. Lysates were mixed v/v with x2 Laemmli sample buffer (Bio-Rad, 161-0737), containing 2-mercaptoethanol (Sigma M7522), and then incubated at 100°C for 5 min. Proteins in non-native conditions were resolved on 10%, 12% or 4-15% Acrylamide/bis gels (161-0173, 161-0175, and 4568084, Bio-Rad).

Then, proteins were transferred to PVDF membranes (170-4159, Bio-Rad) using the trans-blot turbo transfer system (1704150, Bio-Rad) and probed with antibodies for Regnase-1 (Atlas, HPA032053, 1:1000 dilution), β-Actin (Sigma, A5441, 1:50,000 dilution), Flag (Sigma, anti-FLAG M2, F1804, 1:1000 dilution), GFP (Abcam, ab6556, 1:1000 dilution) and IFN-gamma (15365-1-AP, Proteintech, 1:1000 dilution). Band densitometry was done using ImageJ.

### Flow cytometry

Cryopreserved PBMCs were thawed in a water bath at 37°C for approximately three minutes. Cryovial content was transferred to 15 ml tubes and 8 mL of warm (37°C) RPMI 1640 (Gibco) supplemented with 10% Foetal Calf Serum (FCS) (Lonza) 2 mM L-glutamine (Gibco) and 1000 U/mL Penicillin-Streptomycin (Gibco) was slowly added to the sample in the 15 ml tube, while carefully swirling the tube. The cryovials were rinsed with 1 mL of the medium to collect any remaining PBMCs and added to the corresponding 15 ml tubes. Tubes were centrifuged for 10 minutes at 350g at 4°C. After centrifuging, supernatant was removed, the cell pellets were resuspended in cold RPMI. A 10 µl aliquot was taken for each sample and diluted with 10 µl Trypan Blue to stain dead cells. Cells were manually counted using a counting chamber and the number of cells and the cell viability were calculated for each sample. After centrifugation (10 min, 350g, 4°C), supernatant was removed and pellets were diluted in Fluorescence Activated Cell Sorting (FACS) buffer (0.5% Bovine Serum Albumin (BSA) (BSA; Fraction V, Fatty acid free, Calbiochem, San Diego, CA, USA) in Phosphate Buffered Saline (PBS)) to a concentration of at least 500,000 cells/ml with a maximum of 3.5x10^6^ cells/ml, to not exceed the cell concentration suited for the titer of antibodies chosen during the optimization of the panels.

Samples were divided between two panels that were designed to be used in parallel: panel 1 for T cells, unconventional T cells and Innate Lymphoid Cells (ILCs); panel 2 for B cells, monocytes and Dendritic Cells (DCs). Samples were transferred to V-bottom 96-wells plates (Greiner Bio-One), 100 µl of cell suspension per panel and 50-100 µl cell suspension was taken along as unstained control if cell numbers allowed. A reference sample of PBMCs previously isolated from a buffy coat of a healthy donor using density gradient centrifugation with Ficoll-Paque (GE Healthcare) and cryopreserved in aliquots was processed along with the test samples as described above in each experiment, and used later during data quality control (QC) and processing to correct for batch effects.

Plates were centrifuged for 5 minutes at 450g and 4°C, washed once with 150 µl PBS and centrifuged again (5 min, 450g, 4°C). Samples were stained with LIVE/DEAD™ blue fixable viability (Invitrogen) diluted 2000x in PBS, the unstained samples were incubated with PBS only, for 20 minutes. After incubation, 120 µl of FACS buffer was added to all stained and unstained samples and plates were centrifuged (5 min, 450g, room temperature). Then, the samples were stained. First, the staining buffer was added to all stained and unstained samples, immediately followed by the TCR staining mix for panel 1 and IgG for panel 2, after which all plates were incubated for 10 minutes. For panel 1, CXCR5 and CCR7 antibodies were added sequentially and each incubated for 10 minutes before the remaining antibody mix was added, to improve marker resolution. The surface staining antibody mixes were added to the corresponding plates and an equal volume of FACS buffer was added to the unstained samples and incubated for 30 minutes. Incubations were done at room temperature protected from light. After centrifugation (5 min, 450g, room temperature), plates were washed twice with 150 µl FACS buffer.

After surface staining, all samples were fixed and permeabilized by carefully resuspending the cells in 150 µl FoxP3 Fixation/Permeabilization working solution (eBioscience) prepared according to manufacturer’s instructions. After 30 minutes incubation at room temperature in the dark, plates were centrifuged at 800g instead of 450g, to ensure precipitation of the fixed cells. Samples were washed twice with 150 µl 1x Permeabilization buffer (eBioscience) prepared according to manufacturer’s instructions and incubated with 150 µl intracellular staining mix, or Permeabilization buffer only in case of the unstained samples for 45 minutes at room temperature in the dark. Then, samples were washed twice with Permeabilization buffer and centrifuged (5 min, 800g, room temperature). Pellets were resuspended in 75 µl FACS buffer and transferred to mini FACS tubes. Wells were washed with another 75 µl of FACS buffer, which was added to the corresponding FACS tubes. Tubes were sealed with parafilm to prevent evaporation, and stored at 4°C protected from light until data acquisition the next day.

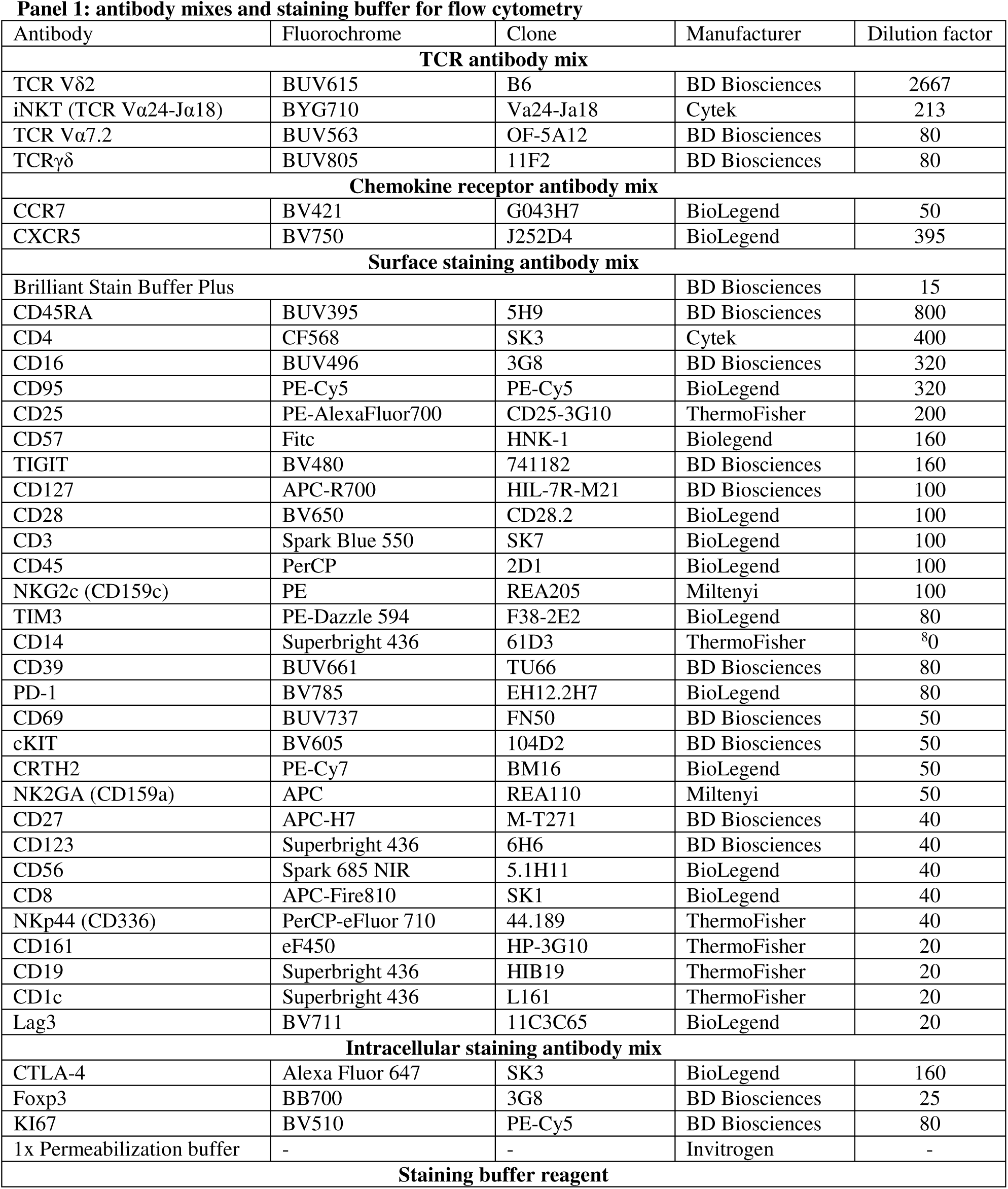
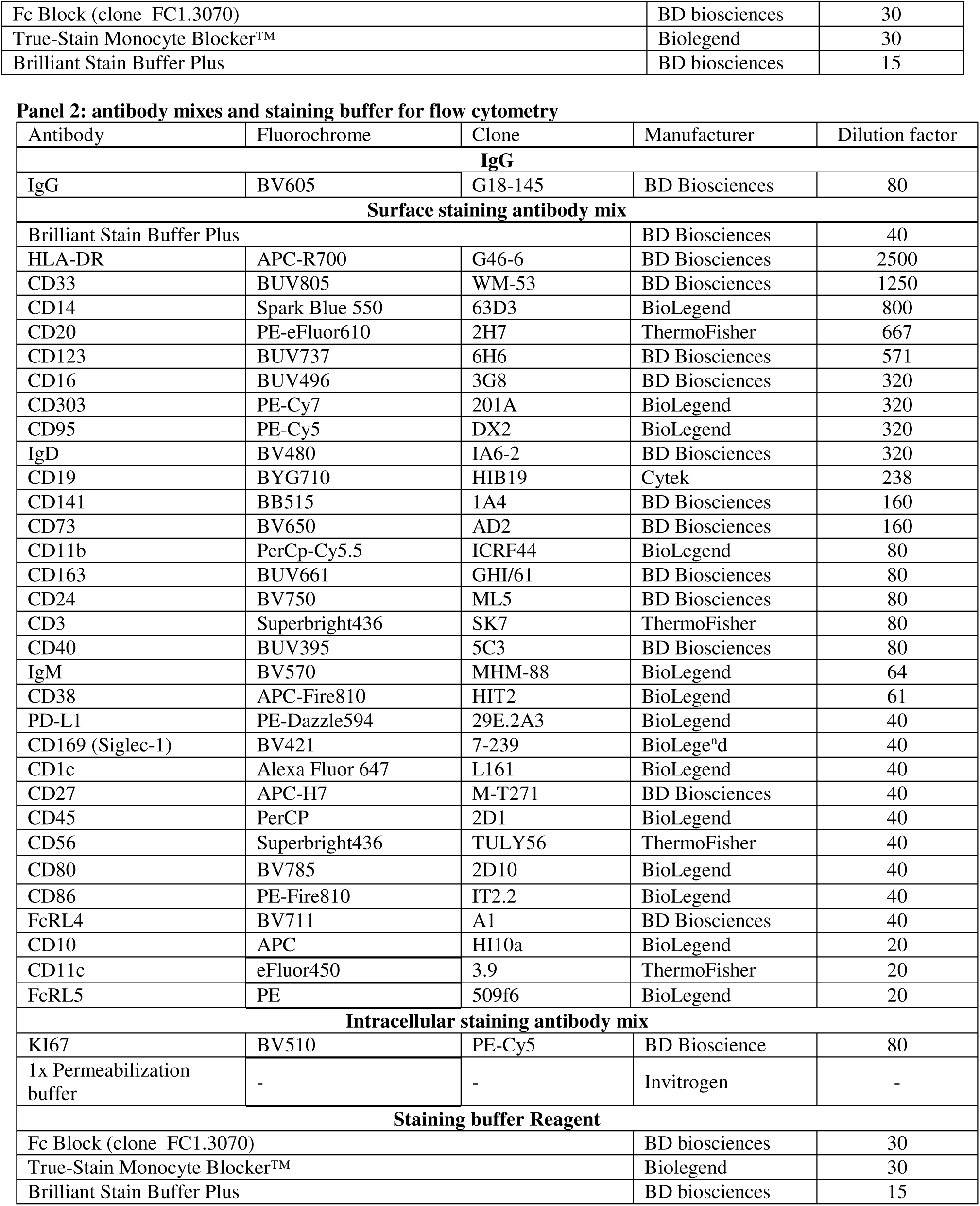

Data were acquired on the Cytek Aurora 5L spectral flow cytometer at high flow rate (60 µl/min). Prior to data acquisition, daily instrument set-up and instrument QC were performed according to the facility guidelines. Single-stained reference controls for each fluorochrome and corresponding antibody in both panels were prepared following the same procedure as the multi-color stained test samples and were run and evaluated prior to acquisition of the test samples. Single stained controls were used for spectral unmixing of the multicolor test samples. The data were unmixed using the SpectroFlo software v2.2.0.4 and minimal compensation was applied when necessary before further data QC and analysis.

### Bulk RNA-seq

PBMCs of patient P1 and two healthy adult controls were studied. PBMCs were plated at 1 million cells/mL and cultured in media containing phytohemagglutinin-L (PHA-L) solution 500X at 2 µl/ml for 5 days. Thereafter, the expanded T cells were washed twice and each sample was divided into 6 (3 conditions x 2 biological replicates each at 10^5^ cells per well). T cells were either left non-stimulated or were stimulated with 1 bead per cell of Dynabeads™ Human T-Activator CD3/CD28 (Gibco) for 4 hours or 24 hours before being collected and lysed in TRIzol (Invitrogen). Total RNA was extracted using RNeasy kit (Qiagen). For controls, 6 RNA samples per subject were sequenced, including 3 conditions x 2 biological replicates. For patient P1, 12 samples were sequenced, including 3 conditions x 2 biological replicates x 2 technical replicates. Sequencing libraries were generated using 1 µg of RNA and KAPA mRNA HyperPrep Kit following manufacturer’s instructions. The library quality was determined on Agilent TapeStation 4200. Sequencing was performed on a NovaSeq 6000 S4 flow cell using v1.5 chemistry (Illumina) generating 25-30 million PE150 reads /sample.

FASTQ sequences were aligned to the human genome build GRCh38 using STAR version 2.7.3a and counted into matrices using FeatureCounts from the Subread Package version 2.0.0. For quality control, the R Package RNAseqQC version 0.1.4 was used with the following sample exclusion criteria: library size less than 5 million counts, clearly dissimilar library complexity compared to other samples, less than 12,500 genes reaching the detection threshold of 10 counts or deviation higher than one log2 fold change when plotted against the mean log2 of the normalized counts of all corresponding replicates. Samples that passed QC were analysed by the R package DESeq2 version 1.36.0. The genes were filtered for a minimum of 5 counts across 2 replicates. Samples were normalized using DESeq2 standard median of ratios method and differential expression was performed between groups of samples with default arguments.

### Single cell RNA-seq

Single cell gene expression libraries of cells from the patient and 2 adult controls, 10,000 PBMCs each, were generated on the 10x Genomics Chromium platform using the Chromium Next GEM Single Cell 3’ Library & Gel Bead Kit v3.1 and Chromium Next GEM Chip G Single Cell Kit (10x Genomics) according to the manufacturer’s protocol. Gene expression libraries were sequenced on a NovaSeq 6000 S4 flow cell using v1.5 chemistry (Illumina) to generate 30,000 reads/cell.

Single-cell RNA sequencing data of age-matched controls were retrieved from a previous study ^8^. Sequencing read files were stored in the EGA repository (EGA accession number EGAD00001007718) in “cram” format. Data access agreement was obtained via the Data Access Committee of the Wellcome Sanger Institute by Dr. Cecilia Domínguez Conde.

Age-matched controls (5 donors) were taken from the healthy cohort^8^ and analysed alongside single-cell data generated from patient and adult control samples as follows. CellRanger pipeline (version 7.0.0, 10x Genomics) was used to perform library demultiplexing, read alignment to the human transcriptome reference (GRCh38-2020-A) and to generate gene expression count matrices. Ambient RNA was subtracted from the raw count matrices generated by Cell Ranger with the *remove-background* module of the CellBender tool (version 0.2.0). Post-CellBender count matrices were then used for downstream analysis with the Scanpy python package (version 1.9.4). Quality control filters at the level of individual cells *(scanpy.pp.filter_cells)* included: minimum of 1000 UMI counts per cell, a minimum of 600 genes per cell and a maximum of 10% mitochondrial genes.

Total counts per cell were normalized to 10,000 (*scanpy.pp.normalize_per_cell*) and the resulting values were log transformed (*sc.pp.log1p)*. Highly variable genes (HVG) selection (*scanpy.pp.highlly_variable_genes)* led to the identification of 2,073 highly variable genes. To exclude variability associated with B and T cell clonality, VDJ gene segments were removed from the HVG set. The resulting 1,894 HVG were used to perform principal component analysis (PCA) using *scanpy.pp.pca.* Data integration was done using Batch-balanced KNN (BBKNN).^31^ We used an approach previously demonstrated for correcting donor-driven effects as well as chemistry-related technical effects (https://github.com/Teichlab/bbknn).^9,32^ We run a first round of integration across samples, using the function *bbknn.bbknn* and donor as batch key and performed a low resolution graph-based leiden clustering. Next we executed the ridge_regression function *(bbknn.ridge_regression)* with chemistry as covariate and low resolution leiden clusters as confounder key. Finally, by using the regressed count matrix, a second round of integration (*bbknn.bbknn)* was performed using donor as batch key.

Leiden graph-based clustering was conducted prior to cell annotation by using *scanp.tl.leiden* (clustering resolution by default). Cell types were first predicted by CellTypist using the Immune_All_Low model (https://www.celltypist.org/models).^9^ Visual inspection of CellTypist predictions and manual refinement of cell annotation was carried out through meticulous evaluation of the expression of known marker genes (fig. S21). In addition, in order to calculate marker genes we performed differential gene expression (DGE) across leiden clusters (https://www.sc-best-practices.org/cellular_structure/annotation.html). The *sc.tl.rank_gene_groups* scanpy function, with Wilcoxon rank sum method was used. To achieve higher resolution, γδ T, Temra, and Treg cells were sub-clustered prior to DGE analysis. The more cluster-specific markers were selected by filtering the genes based on the fraction of cells expressing the gene within and outside the cluster (*scanpy.tl.filter_rank_genes_groups)*.

### Analysis of scRNA-seq datasets of autoimmune diseases

We analysed three published scRNA-seq datasets comprising adult and child patients with systemic lupus erythematosus (SLE) and systemic juvenile idiopathic arthritis (SJIA).^14–16^ The filtered matrices or h5ad files are available at GEO with the following accession numbers: GSE174188, GSE135779, GSE207633. Each dataset was processed as follows. Quality control filters applied included a minimum of 1,000 UMI counts and 600 genes per cell, and a maximum of 10% mitochondrial genes (*scanpy.pp.filter_cells*). Total counts per cell were normalized to 10,000 (*<u>scanpy.pp.normalize_per_cell</u>*), and the resulting values were log-transformed (*sc.pp.log1p*). Expression values of the *VCAM1* and *CD3E* genes per single cell were extracted into a dataframe (adata.to_df(layer="log_transformed")). The total numbers of *VCAM1+CD3E+* cells and their percentages among *CD3E+* cells were calculated for each subject.

