## Supplementary Information for "*VCAM1*-expressing T cells and systemic autoimmunity in Regnase-1 deficiency"

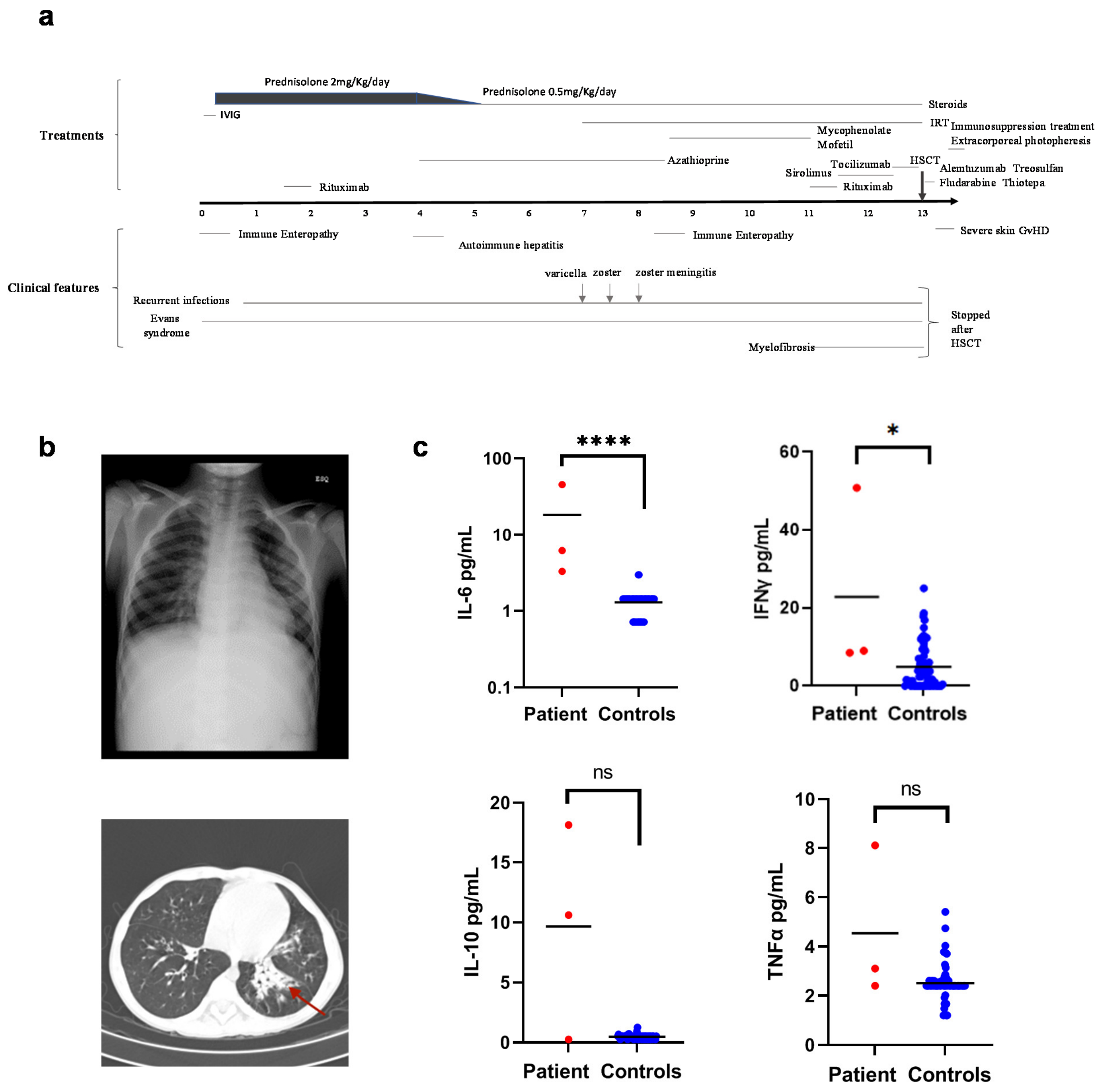


**Supplementary Figure 1. Clinical presentation, serum cytokines and treatment of disease in patient P1**

**(a)** Clinical course and treatment of disease in patient P1. IRT, immunoglobulin replacement therapy; IVIG, intravenous immunoglobulin; HSCT, hematopoietic stem cell transplantation; GvHD, graft versus host disease.

**(b)** Radiological evidence of pneumonia and bronchiectasis in patient P1. Chest X-ray showing interstitial pneumonia (top) and chest CT scan showing bronchiectasis (bottom). Dilated bronchi are shown by red arrow.

**(c)** Serum concentrations of cytokines IL-6, IFNγ, IL-10, and TNFα in patient P1 and healthy controls (N = 65). Mann-Whitney test; * < 0.05, **** < 0.0001; ns, not significant.

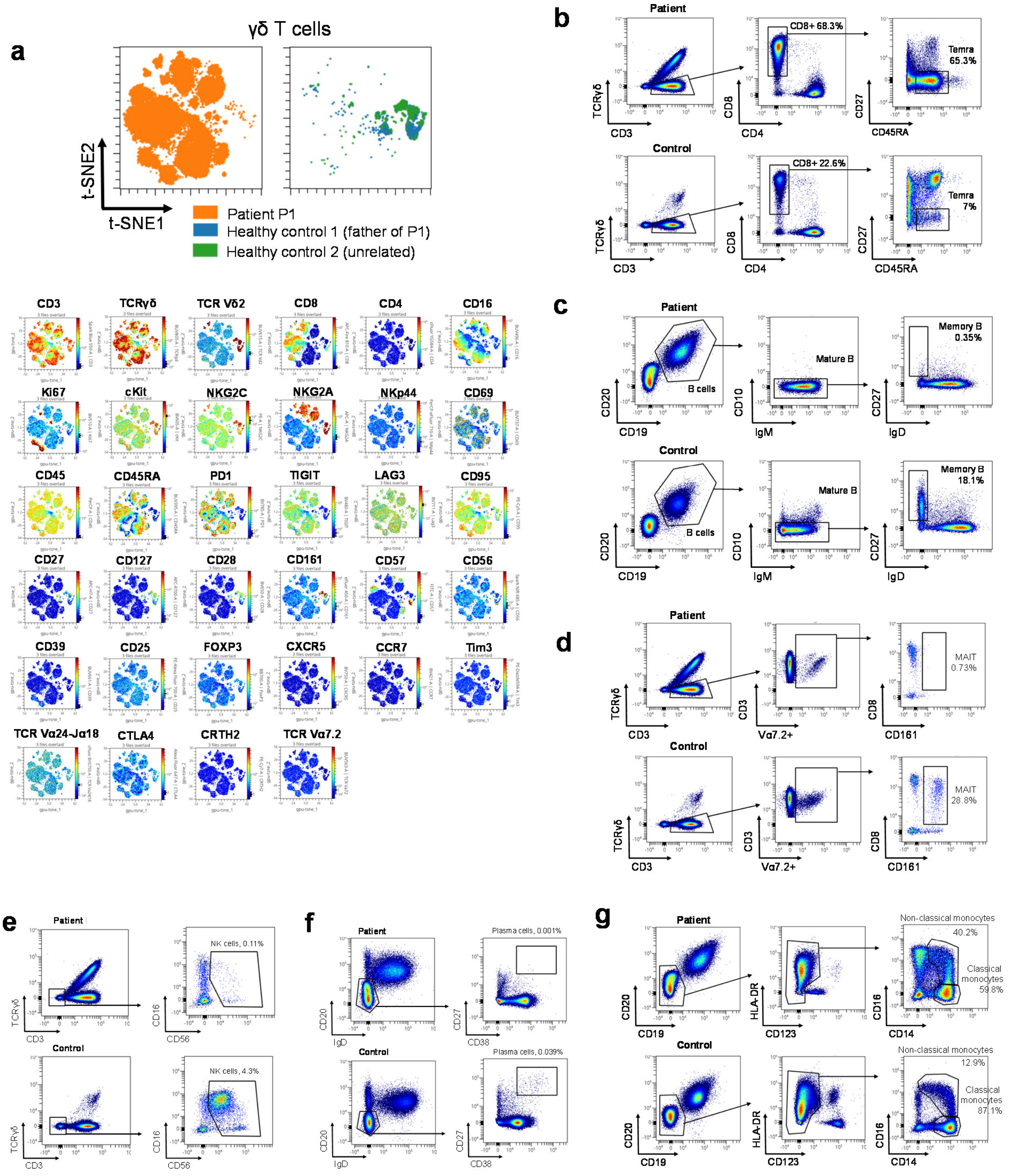


**Supplementary Figure 2. Flow cytometry analysis of PBMCs.**

**(a)** t-SNE analysis of γδ T cells characterised by flow cytometry. (Top) γδ T cells are shown separately in patient P1 and two healthy controls. (Bottom) Marker expression in γδ T cells of patient P1 and two healthy controls.

**(b)** Flow cytometry analysis of CD8+CD27–CD45RA+ terminally differentiated effector memory (Temra) cells in PBMCs. Cell frequencies are shown as % of conventional T cells (middle panel) and as % of CD8+ T cells (right panel).

**(c)** Flow cytometry analysis of CD19+CD20+CD10–CD27+IgD– memory B cells. Frequencies of memory B cells are shown as % of mature B cells.

**(d)** Flow cytometry analysis of CD3+Vα7.2+CD8+CD161+ mucosal-associated invariant T (MAIT) cells. Frequencies of MAIT cells are shown as % of Vα7.2+ T cells.
**(e)** Flow cytometry of NK cells. Frequencies of NK cells are shown as % of total PBMCs.
**(f)** Flow cytometry analysis of CD20–IgD–CD27++CD38++ plasma cells. Frequencies of plasma cells are shown as % of total PBMCs.
**(g)** Flow cytometry analysis of monocytes. Frequencies of classical and non-classical monocytes are shown as % of CD14+ monocytes.

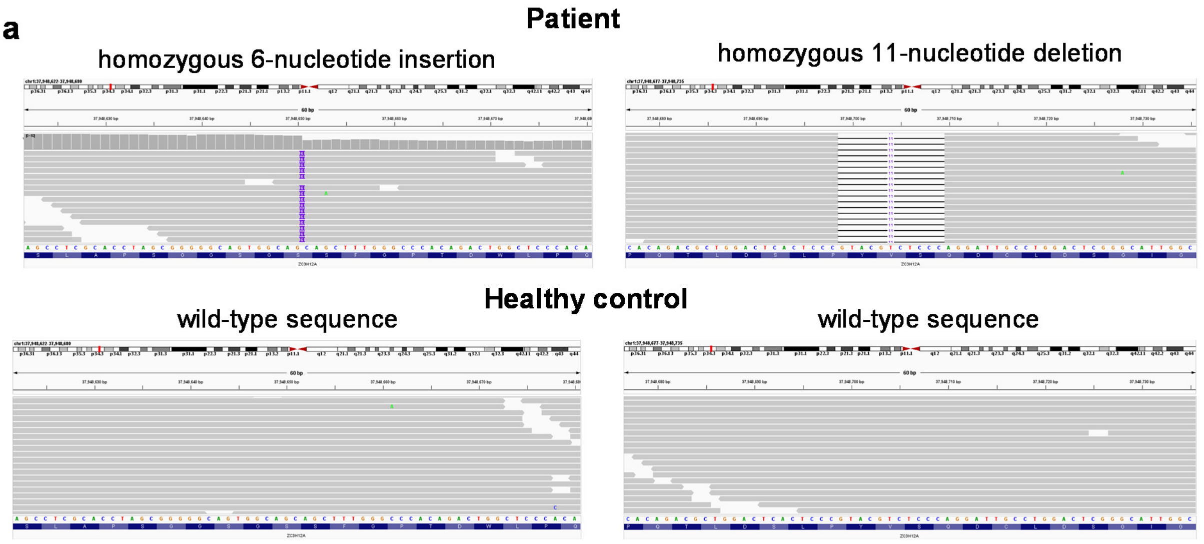


| Chr | Position, hg19 (GRCh38) | Reference/Observed | In gnomAD | Gene (other names) | Ensembl Exon | Nucleotide change | Protein mutation | Frameshift |
|---|---|---|---|---|---|---|---|---|
| 1 | 37,948,650 - 37,948,651 (37,483,049 - 37,483,050) | - / TGGCAG | No | *ZC3H12A* (Regnase-1, MCPIP1) | ENST00000373087 Exon 6 | 6-nucleotide insertion | p.S413_S414insGS | No |
| 1 | 37,948,699 - 37,948,709 (37,483,098 - 37,483,108) | GTACGTCTCCC / - | No | *ZC3H12A* (Regnase-1, MCPIP1) | ENST00000373087 Exon 6 | 11-nucleotide deletion | p.Y430Gfs*27 | Yes |

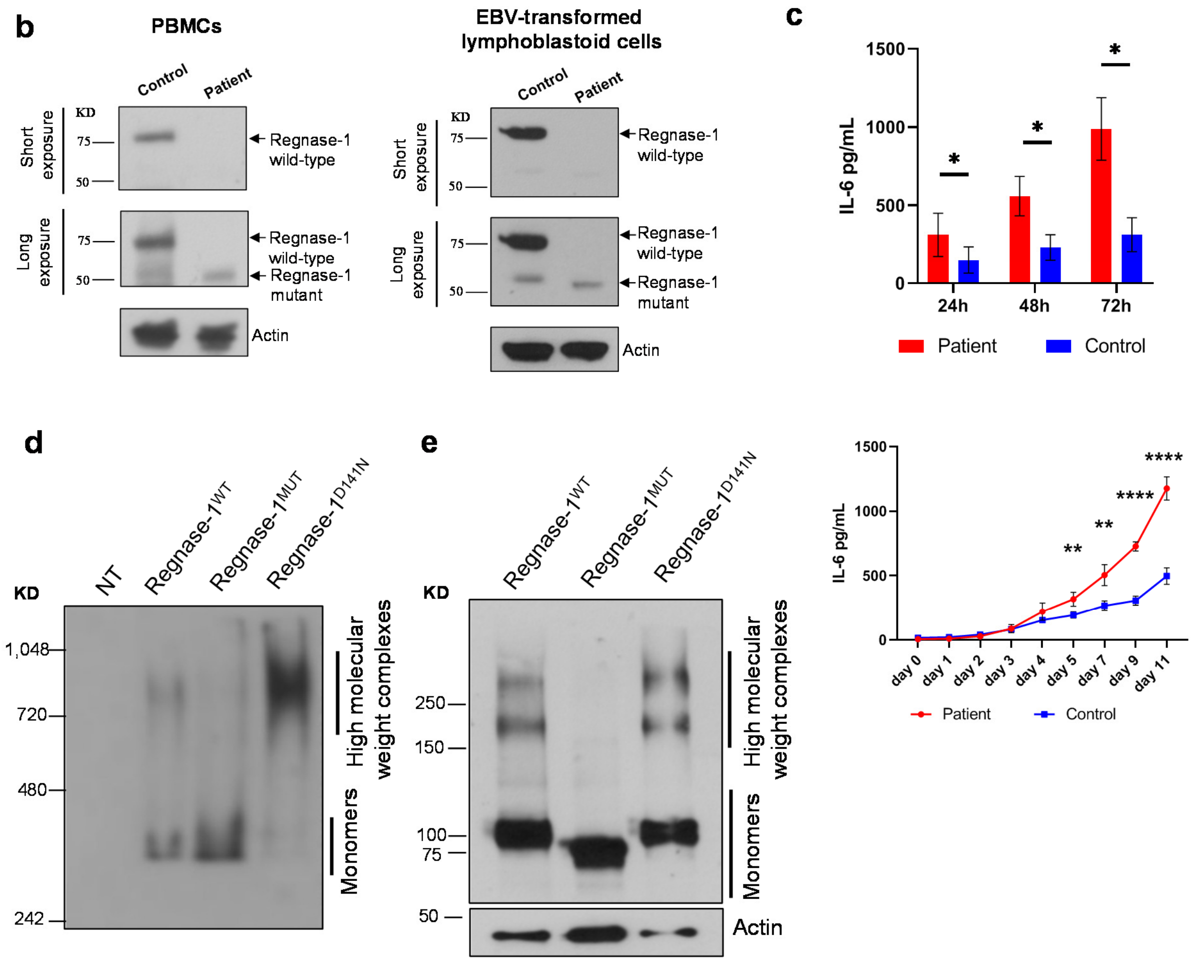


**Supplementary Figure 3. Mutation in the *ZC3H12A* gene leads to reduced expression and impaired oligomerisation of Regnase-1 protein and increased secretion of IL-6.**

(**a**) Homozygous 6-nucleotide insertion and 11-nucleotide deletion in the *ZC3H12A* gene in exome data of patient P1.
(**b**) Immunoblot analysis of the Regnase-1 protein expressed in peripheral blood mononuclear cells (PBMCs) isolated from blood samples, N = 1 (left) and EBV-transformed lymphoblastoid cells, N = 3 (right), representative gel is shown.
(**c**) IL-6 concentrations in supernatants of fibroblasts stimulated with lipopolysaccharide (LPS, 2 µg/mL) (top) and without stimulation (bottom). Top: graph shows concentration values ± SEM, *P*-values were calculated using one-tailed paired t-test (* < 0.05). Bottom: graph shows concentration values ± SD, *P*-values were calculated using one-tailed unpaired t-test (** < 0.01; **** < 0.0001).
(**d**) Lysates of HEK293T cells transfected with plasmids expressing Flag- and GFP-tagged wild-type (Regnase-$1^{WT}$), mutant (Regnase-$1^{MUT}$) or RNase-dead (Regnase-$1^{D141N}$) Regnase-1 proteins were studied using native gel electrophoresis and immunoblotting with an anti-GFP antibody. N=2, representative gel is shown. NT, not transfected.
(**e**) Lysates of HeLa cells lacking endogenous Regnase-1 (HeLa-KO cells) expressing Flag- and GFP-tagged exogenous wild-type (Regnase-$1^{WT}$), mutant (Regnase-$1^{MUT}$) or RNase-dead (Regnase-$1^{D141N}$) Regnase-1 proteins were studied after crosslinking with 1 mM disuccinimidyl suberate (DSS) followed by gel electrophoresis and immunoblotting with an anti-Flag antibody. N=3, representative gel is shown.

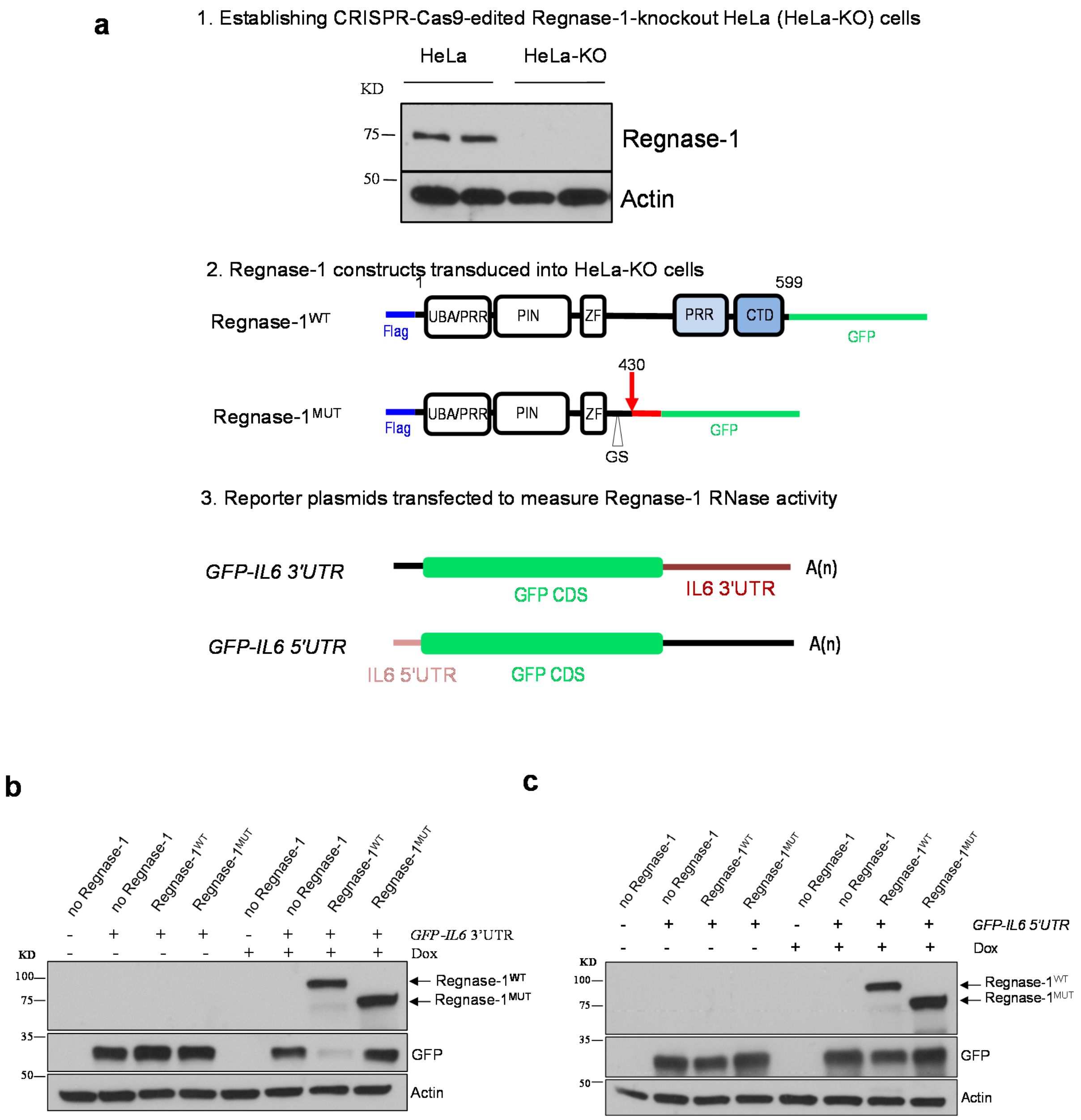


**Supplementary Figure 4. Impaired activity of the mutant Regnase-1 protein.**
**(a)** Experimental system for studying Regnase-1 proteins and measuring their activity. (Top) CRISPR/Cas9 nucleoprotein targeting Regnase-1 was delivered in HeLa cells. Single-cell clones were expanded. Regnase-1 expression was tested by immunoblotting and a Regnase-1-knockout HeLa cell clone (HeLa-KO) was identified. (Middle) HeLa-KO cells were transduced with constructs encoding Regnase-$1^{WT}$ or Regnase-$1^{MUT}$ proteins tagged with Flag on the N terminus and green fluorescent protein (GFP) on the C terminus. Regnase-$1^{MUT}$ has mutation found in patient P1, including insertion of 2 amino acids (p.S413_S414insGS) and a frameshift deletion that affects Regnase-1 protein at position 430 (p.Y430Gfs*27, shown by red arrow); the premature Stop codon in Regnase-$1^{MUT}$ has been removed to allow GFP expression. Regnase-1 protein domains and motifs: ubiquitin-associated domain (UBA), PilT N-terminus nuclease (PIN) responsible for RNase activity, zinc-finger motif (ZF), proline-rich region (PRR), C-terminal domain (CTD). (Bottom) Reporter plasmids encoding GFP coding sequence (CDS) followed by the *IL6* 3′UTR (a known Regnase-1 target) or preceded by *IL6* 5′UTR (a negative

control containing sequence not targeted by Regnase-1) were transfected in HeLa-KO cells. The bottom panel is adapted from Hutin et al.[30]

**(b)** Regnase-1 activity assay: Regnase-1-knockout HeLa (HeLa-KO) cells either not expressing Regnase-1 or expressing Flag- and GFP-tagged exogenous Regnase-$1^{WT}$ and Regnase-$1^{MUT}$ proteins under the control of the Dox-inducible promoter were transfected with plasmid *GFP-IL6 3'UTR*. +Dox indicates overnight incubation with 20 ng/mL doxycycline. Representative immunoblot shows Regnase-$1^{WT}$, Regnase-$1^{MUT}$ and GFP expression, and actin as a loading control. N = 4. GFP densitometry results are shown in Figure 1i.

**(c)** Analysis of activity of Regnase-$1^{WT}$ and Regnase-$1^{MUT}$ proteins expressed in HeLa-KO cells as in (B), but transfected with a plasmid expressing *GFP-IL6 5'UTR* (negative control). N = 2.

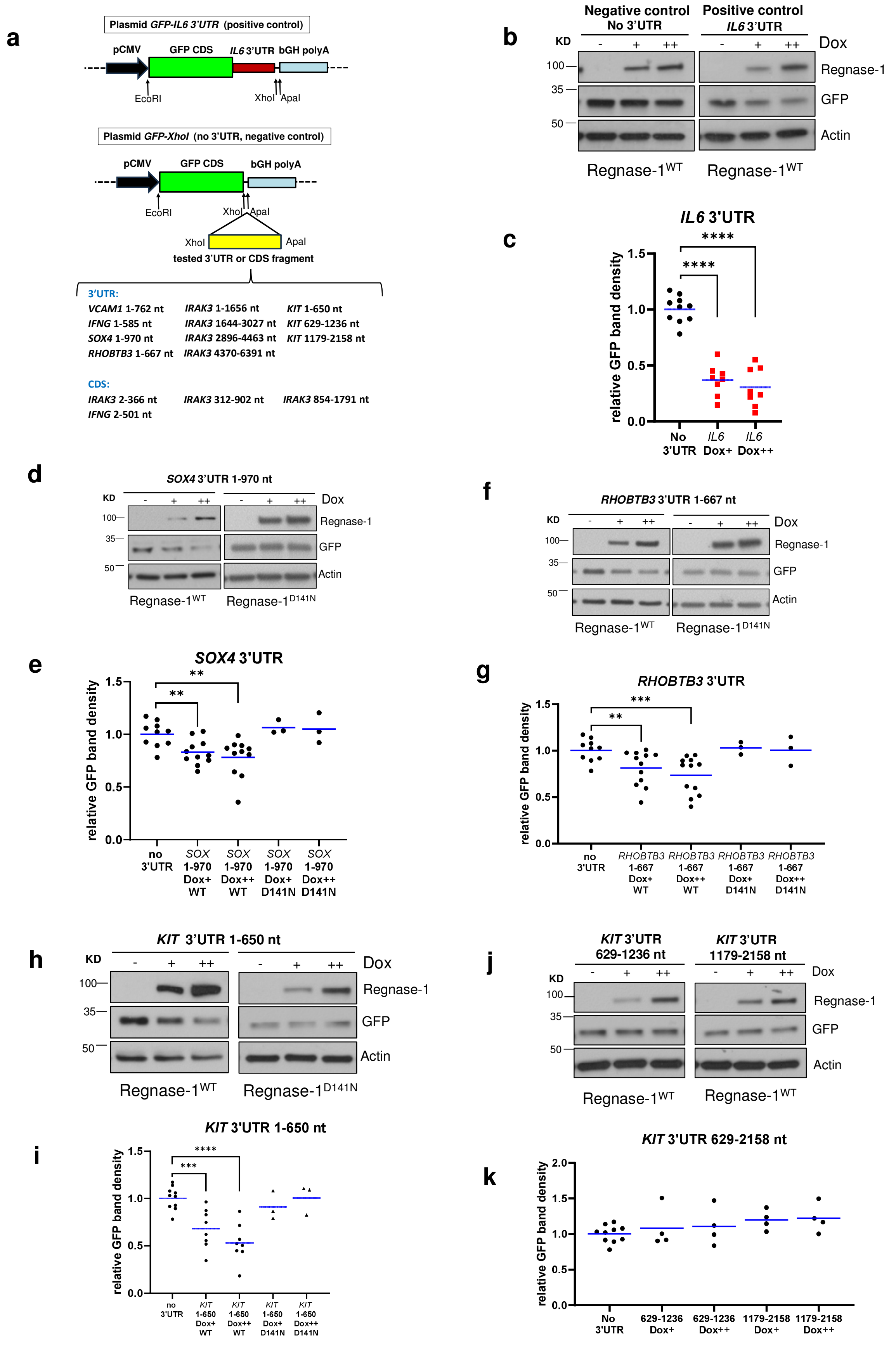


**Supplementary Figure 5. 3′UTRs of the *SOX4, RHOBTB3,* and *KIT* mRNAs are directly targeted by Regnase-1.**

**(a)** Assay schematic: GFP-expressing plasmid *GFP-IL6 3'UTR* was used as a positive control. To engineer a negative control (no 3′UTR), the IL6 3′UTR was removed. Then, 3′UTRs or coding sequences (CDS) of candidate mRNAs were cloned in the GFP-expressing plasmid instead of the IL6 3′UTR.
**(b, d, f, h, j)** Regnase-1activity assay: Regnase-1-knockout HeLa (HeLa-KO) cells expressing exogenous wild-type Regnase-1 (Regnase-$1^{WT}$) under the control of the doxycycline (Dox)-inducible promoter were transfected with a plasmid encoding GFP with no 3′UTR (negative control) or GFP followed by *IL6* 3′ UTR (positive control) or a candidate fragment. Cells were incubated overnight either without doxycycline (Dox–) or with doxycycline at final concentrations of 10 ng/mL (Dox+) and 20 ng/mL (Dox++). $N \geq 3$, representative immunoblots are shown.
**(c, e, g, i, k)** Band densitometry: the graphs show raw integrated density of the GFP band in Dox+ and Dox++ relative to Dox– corrected for the background and actin band density. Each dot represents a separate transfection experiment. Means are shown by blue lines. WT, Regnase-$1^{WT}$. *P*-values were calculated using one-tailed Mann-Whitney test; ** $< 0.01$, *** $< 0.001$, **** $< 0.0001$.

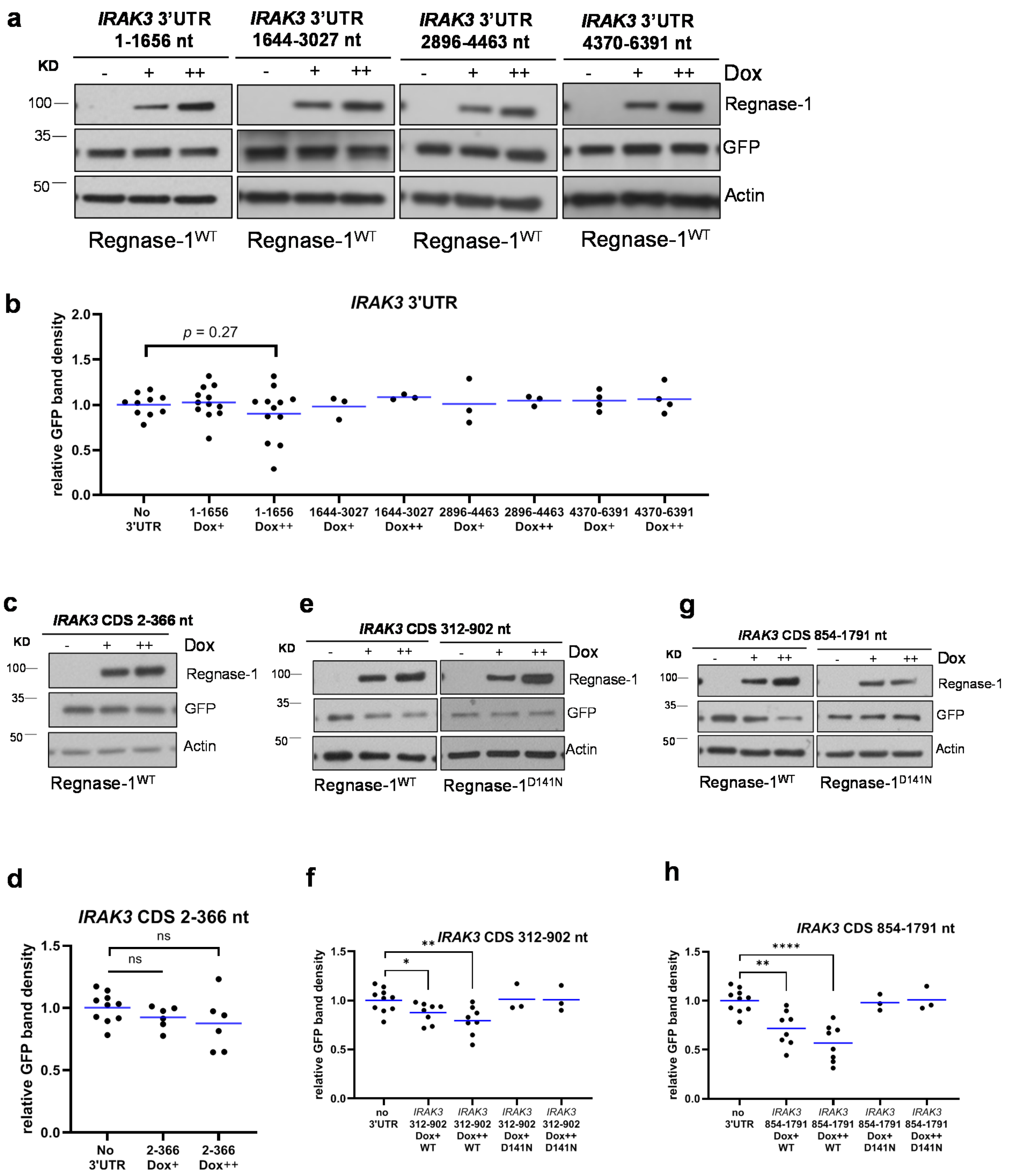


**Supplementary Figure 6. Regnase-1 targets the coding sequence (CDS) of the *IRAK3* mRNA but not its 3′UTR.**

**(a, c, e, g)** Regnase-1 activity assay: HeLa-KO cells expressing exogenous Regnase-1$^{WT}$ under the control of the doxycycline (Dox)-inducible promoter were transfected with a plasmid encoding GFP with no 3′UTR (negative control) or a candidate 3′UTR or CDS fragment. Cells were incubated overnight either without doxycycline (Dox–) or with doxycycline at final concentrations of 10 ng/mL (Dox+) and 20 ng/mL (Dox++). N ≥ 3, representative immunoblots are shown.

**(b, d, f, h)** Band densitometry: the graphs show raw integrated density of the GFP band in Dox+ and Dox++ relative to Dox– corrected for the background and actin band density. Each dot represents a separate transfection experiment. Means are shown by blue lines. *P*-values were calculated using one-tailed Mann-Whitney test. * < 0.05, ** < 0.01, **** < 0.0001; ns, not significant.

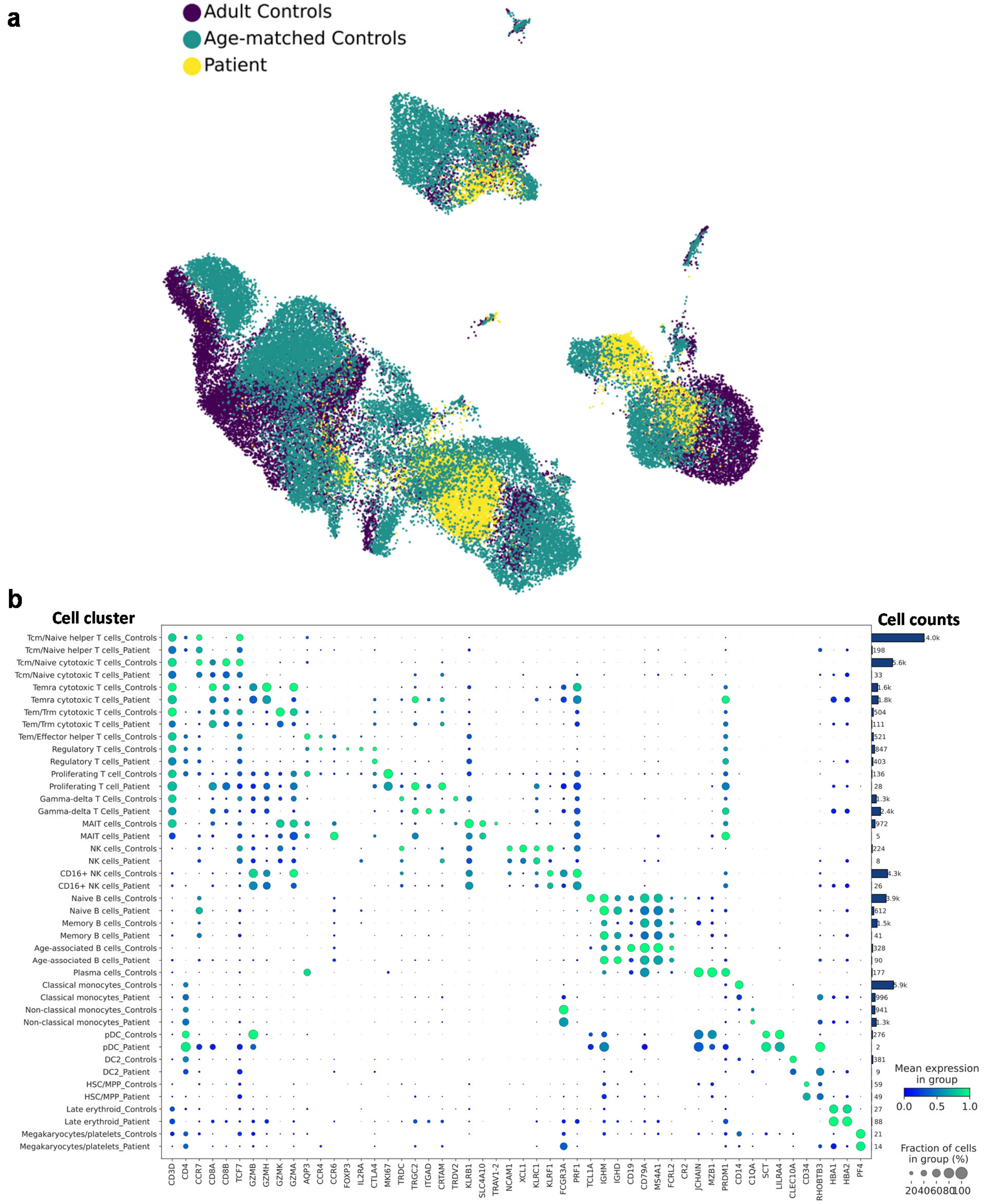


**Supplementary Figure 7. scRNA-seq analysis of PBMCs.**
**(a)** Uniform Manifold Approximation and Projection (UMAP) of the single cell RNA-seq (scRNA-seq) analysis of PBMCs from patient P1 and 7 healthy controls, including 5 age-matched children (NP30, NP31, NP39, NP41 and NP44) from Yoshida et al[8] and 2 adults. **(b)** Dotplot showing gene expression of cell lineage marker genes.

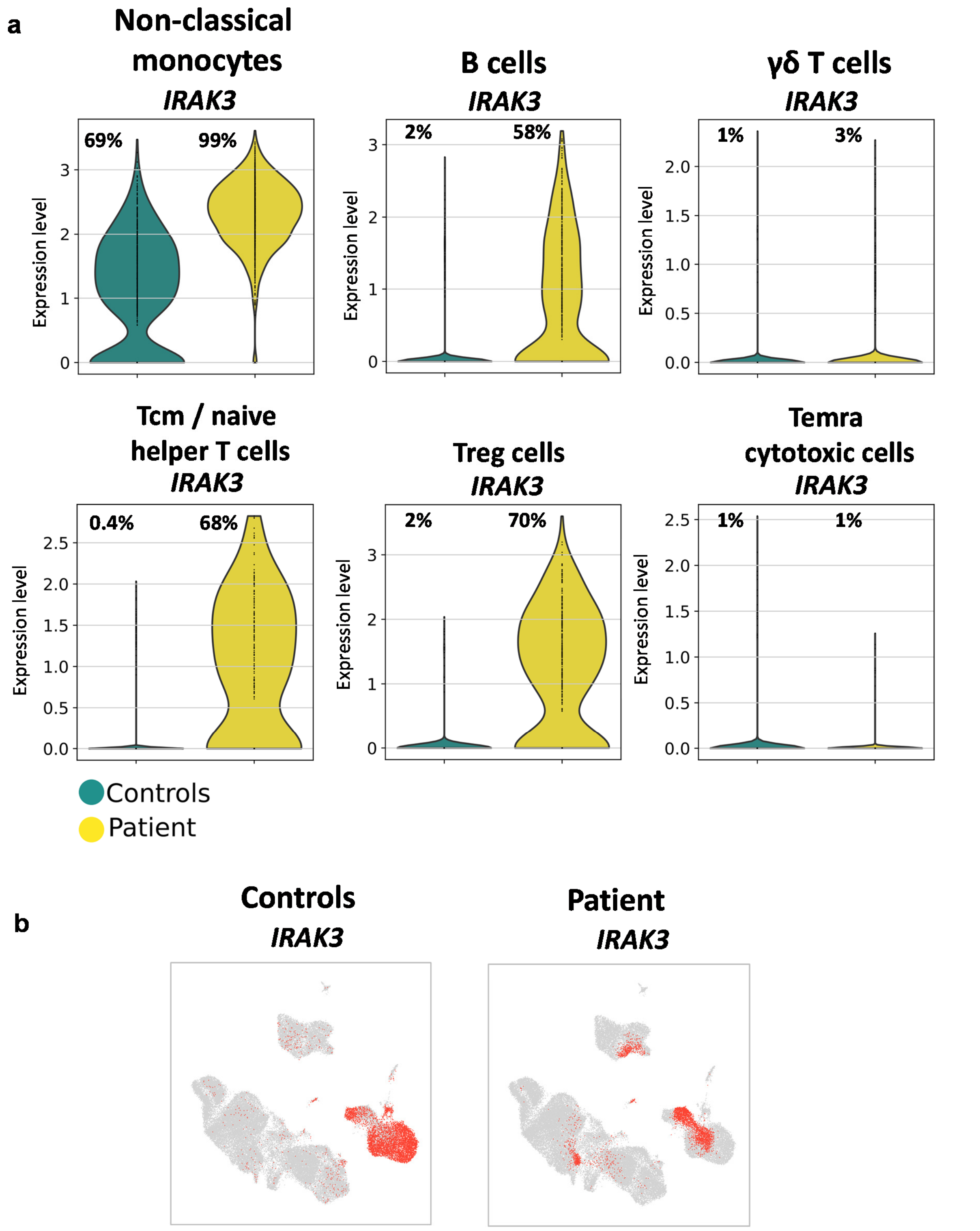


**Supplementary Figure 8.** ***IRAK3*** **is expressed in non-classical monocytes, B and T cells of patient P1.**
**(a)** Violin plots show *IRAK3*-expressing cells. Percentages of cells with gene expression level > 0 are shown on each plot. B cells correspond to clusters 12-14.
**(b)** UMAP plots show in red *IRAK3*-expressing cells (expression level > 0) of 7 controls and patient P1 respectively; cells where gene expression level = 0 are shown in grey.

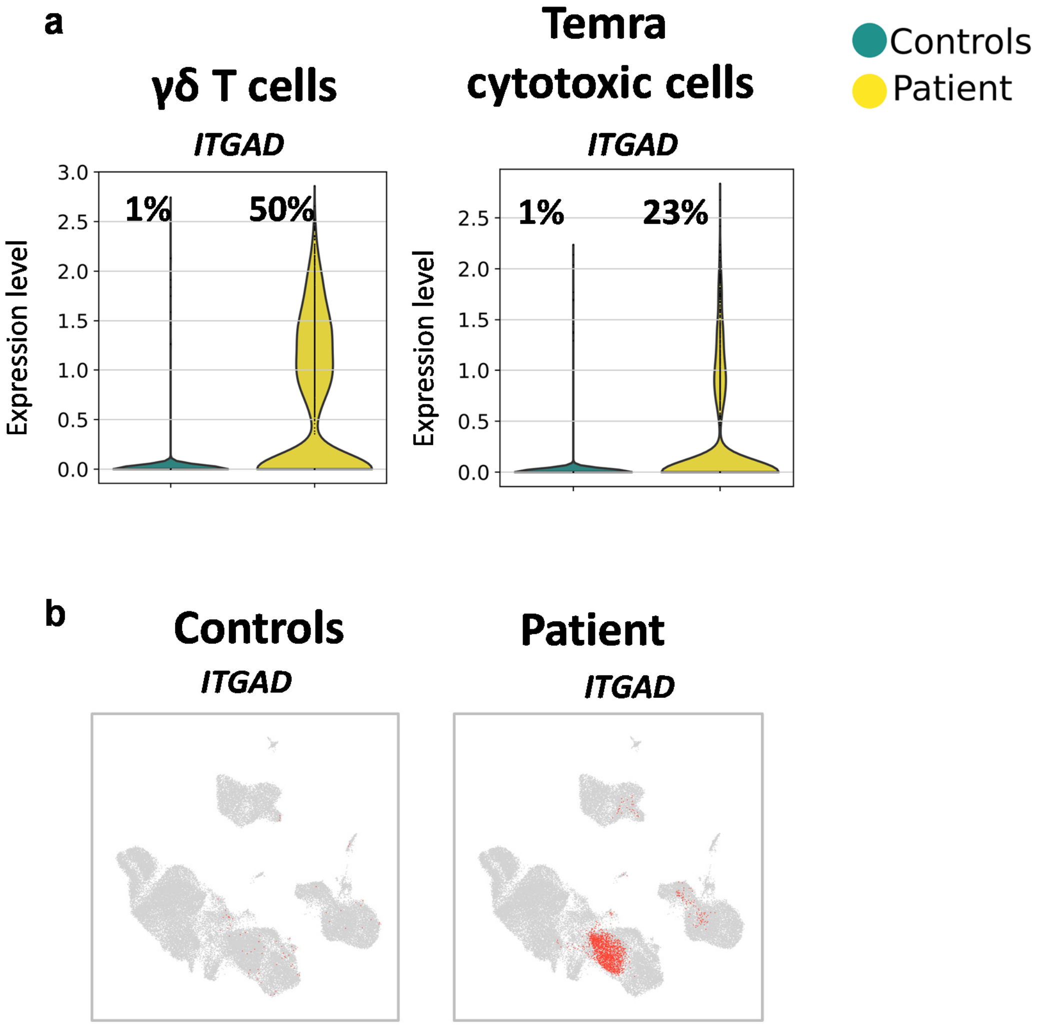


**Supplementary Figure 9.** ***ITGAD*** **is expressed in γδ T cells and Temra cytotoxic cells of patient P1.**

**(a)** Violin plots show *ITGAD*-expressing cells. Percentages of cells with gene expression level > 0 are shown on each plot.

**(b)** UMAP plots show in red *ITGAD* -expressing cells (expression level > 0) of 7 controls and patient P1 respectively; cells where gene expression level = 0 are shown in grey.

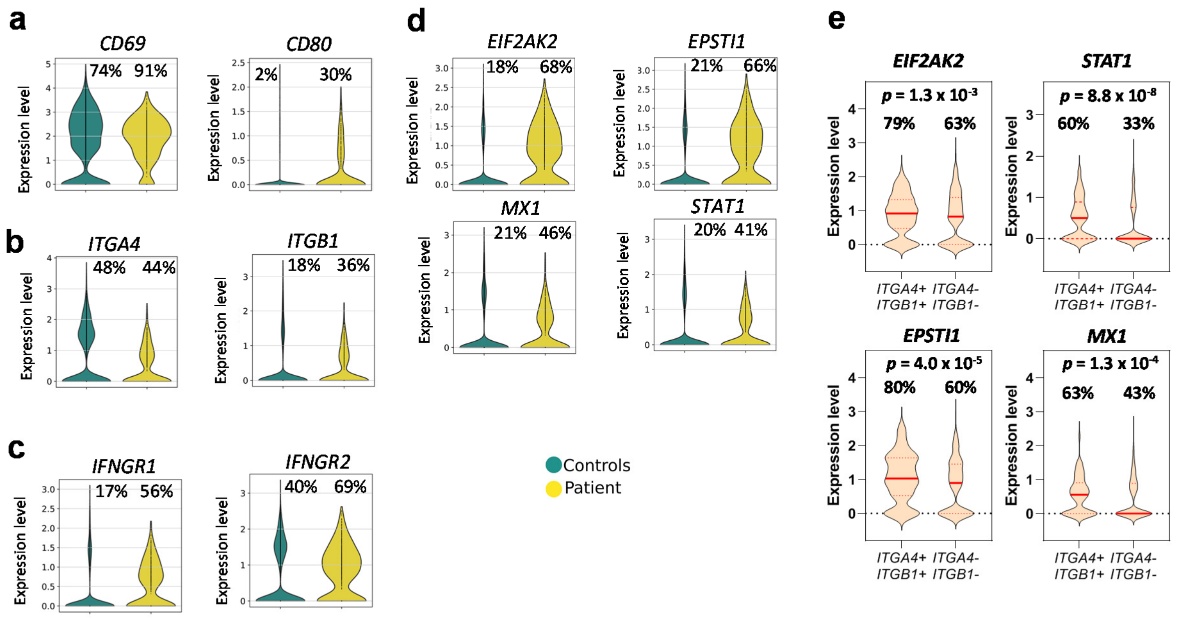


**Supplementary Figure 10. B cells of patient P1 express genes encoding activation markers, integrin α4β1, IFNγ receptor and IFN-response genes.**
**(a, b, c, d)** Violin plots show gene-expressing cells. Percentages of cells with gene expression level > 0 are shown on each plot. B cells correspond to clusters 12-14.
**(e)** Violin plots showing expression of IFN-response genes in B cells (clusters 12-14) categorised by the expression of genes encoding integrin α4β1 in the same cells. Percentages of cells with gene expression level > 0 are shown on each plot. Median is shown by the thick red line, quartiles are shown by dot red lines. *P*-values are from two-tailed chi-square tests with 1 df.

**Supplementary Table 1. Immunological and biochemical analyses of blood in patient P1.**

| | Initial analysis | Age 2 – 6 years | | | | Age 7 – 12.5 years | | | | | | | Control range |
|---|---|---|---|---|---|---|---|---|---|---|---|---|---|
| Haemoglobin (g/L) | **5** | 12 | **10.7** | **10.9** | **9.6** | 13.1 | **7.4** | 12.6 | 11.7 | **8.9** | **5.1** | **7** | 11.5 - 13.5 |
| Reticulocyte count (%) | **20** | **3** | **3.9** | 1.84 | **11.7** | | **10** | | | | **0.3** | **5** | 0.5 - 2.5 |
| Platelets (x $10^9$/L) | **40** | 250 | **100** | 576 | 480 | 354 | 275 | **181** | **157** | **120** | **60** | **150** | 200 - 450 |
| Leukocytes (cells/µL) | **15,150** | 10,570 | **14,150** | 11,280 | 10,700 | 10,900 | 9,460 | 10,100 | **14,000** | 13,000 | 4,660 | 7,090 | 4,500 - 13,000 |
| Neutrophils | **9,530** | 5,790 | 7,530 | 6,410 | 6,500 | 8,100 | 7,460 | 6,060 | **10,640** | **11,440** | 3,360 | 3,300 | 1,500 - 8,500 |
| Monocytes | 350 | 450 | 940 | 610 | 900 | **1,100** | 460 | 1310 | 780 | **170** | **160** | 370 | 200 - 1,000 |
| Basophils | 70 | 100 | 60 | 80 | 100 | 0 | 40 | 0 | 40 | 10 | 20 | 0 | 0 - 100 |
| Eosinophils | 450 | 480 | **1,070** | 830 | 100 | 400 | 140 | **30** | **10** | **0** | **0** | **0** | 100 - 800 |
| Lymphocytes (cells/µL) | 4,750 | 3,750 | 4,500 | 3,200 | 3,100 | **1,400** | **1,400** | 2,620 | 2,530 | **1,380** | **1,120** | 3,410 | 2,500 - 5,500 |
| T cells | | | | | | | | | | | | | |
| CD3+ (cells/µL) | | | 2,700 | 2,150 | | | | | | **3,317** | | | 1,200 - 3,000 |
| CD3+ CD4+ (cells/µL) | | | 1,430 | 1,200 | | | | | | 763 | | | 650 - 1,500 |
| CD3+ CD4+ CD45RA+ (%) | | | 65 | 60 | | | | | | **26** | | | 53 - 86% |
| CD3+ CD8+ (cells/µL) | | | **1,200** | 800 | | | | | | **2,146** | | | 370 - 1,100 |
| CD3+ CD8+ CD45RA+ (%) | | | 67 | 62 | | | | | | **7** | | | 42 - 82% |
| CD3+ CD4- CD8- TCR αβ (%) | | | | **3** | | | | | | | | | <2% |
| NK cells | | | | | | | | | | | | | |
| CD16+ CD56+ (cells/µL) | | | 180 | 140 | | | | | | 132 | | | 100 - 480 |
| B cells | | | | | | | | | | | | | |
| CD19+ (cells/µL) | | | | 341 | | | 300 | | | 480 | | | 270 - 860 |
| CD19+ CD27- IgD+ IgM+ (%) | | | | | | | **84** | | | **85** | | | 47 - 70% |
| CD19+ CD27+ (%) | | | | | | | 11 | | | 8 | | | 7 - 24% |
| CD19+ CD27+ IgD- (class-switched) | | | | | | | 3 | | | 5 | | | 2.7 - 12.5% |
| CD19+ CD38+ $IgM^{high}$ (transitional) | | | | | | | 8 | | | 0 | | | |
| Immunoglobulins | | | | | | | | | | | | | |
| IgA (g/L) | **0** | | **0** | **0** | **0** | **0** | **0** | **0** | **0** | **0** | **0** | | 0.2 - 2.5 |
| IgG (g/L) | **2.3** | | **32.0** | **20.3** | **23.7** | **17.0** | 10.0 | 9.3 | 7.5 | 6.5 | 8.7 | | 4.2 - 14.8 |
| IgM (g/L) | 2.2 | | **2.7** | **4.6** | **5.8** | **5.4** | **3.4** | **5.3** | **5.7** | **9.8** | **5.0** | | 0.5 - 2.2 |
| IgE (IU/mL) | **45** | | | | **15** | | | 9.5 | 0 | 0 | 0 | | <10 |

| | | | | | | | | | | | | | |
|---|---|---|---|---|---|---|---|---|---|---|---|---|---|
| Autoantibodies | | | | | | | | | | | | | |
| Coombs test | **IgG+++** | **IgG+** | **IgG++** | **IgG+** | **IgG+** | **IgG+** | **IgG+++** | **IgG+** | | | Neg. | Neg. | Neg. |
| | **C3d+++** | **C3d+** | **C3d+** | **C3d+** | **C3d+** | **C3d+** | **C3d+++** | **C3d+** | **C3d+** | **C3d+** | | | Neg. |
| Anti-transglutaminase | 0.08 | 0.04 | | | | | 0 | | | 0 | | | < 20 |
| Anti-enterocyte | Neg. | Neg. | | | | | | | | | | | Neg. |
| Anti-LKM | | | **Positive** | **Positive** | Neg. | | Neg. | | Neg. | Neg. | | | Neg. |
| Anti-dsDNA (UI/mL) | | | 12.9 | **30** | 15 | 10 | 12 | | | 0 | | | < 20 |
| ASMA | | | **Positive** | **Positive** | **Positive** | Neg. | **Positive** | Neg. | Neg. | Neg. | | | Neg. |
| AMA | | | Neg. | | | | Neg. | Neg. | | Neg. | | | Neg. |
| ANA | **1/160** | **1/160** | | | | | **1/160** | **1/160** | | **1/160** | | | Neg. |
| Anti-thyroglobulin | | | Neg. | | | Neg. | Neg. | Neg. | | Neg. | | | Neg. |
| Anti-TPO | | | Neg. | | | Neg. | Neg. | Neg. | | Neg. | | | Neg. |
| Anti-gliadin | Neg. | Neg. | | | | | Neg. | | | Neg. | | | Neg. |
| Anti-Diphtheria (IU/mL) | | | | | | **0** | 0.6 | | | **0.18** | | | > 0.5 |
| Anti-Tetanus (IU/mL) | | | | | | **0** | 3.5 | | | **0** | | | > 0.5 |
| Anti-Pneumococcus | | | | | | | | **19** | **22** | | | | > 25 |
| Biochemical analysis | | | | | | | | | | | | | |
| Haptoglobin (g/L) | <0.07 | 0.2 | <0.07 | 0.35 | 0 | | | | | | | | 0.34 - 2 |
| AST (U/L) | 25 | 30 | **200** | 18 | 20 | 27 | 35 | 27 | 30 | **40** | **274** | **40** | 8 - 35 |
| ALT (U/L) | 20 | 20 | **171** | 10 | 11 | 17 | 20 | 22 | 20 | 15 | **72** | 27 | 0 - 55 |
| GGT (U/L) | 40 | | **131** | | | 40 | | 25 | | | **137** | 25 | < 55 |
| Alkaline phosphatase (U/L) | 250 | | 444 | | | 300 | 355 | | 280 | 300 | **880** | 320 | < 500 |
| Total bilirubin (mg/dL) | 10.8 | | 0.71 | 0.9 | 0.8 | 0.7 | 1.2 | 0.8 | 0.9 | **1.4** | **2.02** | **1.4** | 0.3 - 1.2 |
| Direct bilirubin (mg/dL) | **0.9** | | 0.3 | 0.2 | 0.2 | 0.2 | 0.2 | 0.4 | 0.2 | 0.4 | 0.4 | 0.2 | 0 - 0.5 |
| Calprotectin (ug/g) | **850** | <50 | | | | 20 | 30 | | **600** | **550** | **2,800** | **700** | < 50 |

Abnormal results are shown in bold. ANA, anti-nuclear antibody; anti-LKM, anti-kidney liver microsome antibody; ASMA, anti-smooth muscle antibody; AMA, anti-mitochondrial antibody; anti-TPO, anti-thyroid peroxidase antibody. AST, aspartate transaminase; ALT, alanine transaminase; GGT, gamma-glutamyl transferase; Neg., negative.

**Supplementary Table 2. Flow cytometry analysis of PBMCs in patient P1 and 2 healthy controls.**

| | | | Patient P1 | | Control (unrelated) | | Control (father) | |
|---|---|---|---|---|---|---|---|---|
| **Cell type** | **Markers** | **Cells** | **n** | **%** | **n** | **%** | **n** | **%** |
| *Marker panel 1* | *singlets live CD45+* | *total cells* | *200,000* | | *200,000* | | *200,000* | |
| iNKT | Vα24-Jα18+ Vα7.2- | (% of total cells) | 12 | 0.006 | 109 | 0.055 | 15 | 0.008 |
| **NK cells** | **Vα24-Jα18- TCRγδ- CD3- CD56+** | **(% of total cells)** | **229** | **0.11** | **8,583** | **4.3** | **12,302** | **6.2** |
| CD16+ NK | $CD56^{dim}$ CD16+ | (% of NK) | 126 | 55.0 | 7,982 | 93.0 | 11,354 | 92.3 |
| CD16- NK | $CD56^{bright}$ CD16- | (% of NK) | 102 | 44.5 | 589 | 6.9 | 923 | 7.5 |
| ILCs | Vα24-Jα18- TCRγδ- CD3- CD16- CD56- CD127+ | (% of total cells) | 266 | 0.13 | 184 | 0.092 | 122 | 0.061 |
| ILC1 | CRTH2- cKit- | (% of ILCs) | 188 | 70.7 | 59 | 32.1 | 33 | 27.0 |
| ILC2 | CRTH2+ | (% of ILCs) | 35 | 13.2 | 75 | 40.8 | 41 | 33.6 |
| ILC3 | CRTH2- cKit+ | (% of ILCs) | 37 | 13.9 | 45 | 24.5 | 47 | 38.5 |
| **γδ T cells** | **CD3+ TCRγδ+** | **(% of total cells)** | **42,768** | **21.4** | **905** | **0.45** | **3,906** | **2.0** |
| Vδ2 T cells | Vδ2+ | (% of total γδ T cells) | 1,292 | 3.0 | 664 | 73.4 | 3,048 | 78.0 |
| **Vδ1 T cells** | **Vδ2-** | **(% of total γδ T cells)** | **41,461** | **96.9** | **239** | **26.4** | **857** | **21.9** |
| Vδ1 T CD8+ PD1+ | CD8+ PD1+ | (% of Vδ1 γδ T cells) | 20,550 | 49.6 | 24 | 10.0 | 87 | 10.2 |
| Vδ1 T CD8+ PD1- | CD8+ PD1- | (% of Vδ1 γδ T cells) | 1,853 | 4.5 | 74 | 31.0 | 73 | 8.5 |
| Vδ1 T CD8- PD1+ | CD8- PD1+ | (% of Vδ1 γδ T cells) | 17,478 | 42.2 | 34 | 14.2 | 209 | 24.4 |
| Vδ1 T CD8- PD1- | CD8- PD1- | (% of Vδ1 γδ T cells) | 1,580 | 3.8 | 107 | 44.8 | 488 | 56.9 |
| Vα7.2 T cells | CD3+ TCRγδ- Vα7.2+ | (% of total cells) | 1,506 | 0.75 | 3,057 | 1.53 | 2,516 | 1.26 |
| **MAIT** | **CD8+ CD161+** | **(% of Va7.2+ T cells)** | **11** | **0.73** | **879** | **28.8** | **582** | **23.1** |
| CD161- MAIT | CD8+ CD161- | (% of Va7.2+ T cells) | 1,099 | 73.0 | 891 | 29.1 | 1,234 | 49.0 |
| CD8- MAIT | CD8- CD161+ | (% of Va7.2+ T cells) | 34 | 2.3 | 279 | 9.1 | 118 | 4.7 |
| CD4+ MAIT | CD4+ | (% of Va7.2+ T cells) | 277 | 18.4 | 941 | 30.8 | 529 | 21.0 |
| Conventional T cells | CD3+ TCRγδ- Vα7.2- | (% of total cells) | 104,726 | 52.4 | 96,346 | 48.2 | 100,136 | 50.1 |
| DP T cells | CD4+ CD8+ | (% of conventional T cells) | 1,022 | 1.0 | 410 | 0.4 | 1,709 | 1.7 |
| DN T cells | CD4- CD8- | (% of conventional T cells) | 707 | 0.7 | 1,030 | 1.1 | 971 | 1.0 |
| **CD8+ T cells** | **CD4- CD8+** | **(% of conventional T cells)** | **71,505** | **68.3** | **21,772** | **22.6** | **26,877** | **26.8** |
| **CD8+ naive** | **CD27+ CD45RA+ CD95-** | **(% of CD8+ T cells)** | **389** | **0.54** | **8,430** | **38.7** | **2,794** | **10.4** |
| CD8+ Tscm | CD27+ CD45RA+ CD95+ | (% of CD8+ T cells) | 375 | 0.52 | 481 | 2.2 | 4,011 | 14.9 |
| **CD8+ Temra** | **CD27- CD45RA+** | **(% of CD8+ T cells)** | **46,717** | **65.3** | **1,533** | **7.0** | **1,861** | **6.9** |
| CD8+ Tem | CD45RA- | (% of CD8+ T cells) | 19,951 | 27.9 | 9,446 | 43.4 | 12,545 | 46.7 |
| **CD8+ Tcm** | **CD45RA- CCR7+ CD28+** | **(% of CD8+ T cells)** | **893** | **1.2** | **4,362** | **20.0** | **7,170** | **26.7** |

| | | | | | | | | |
|---|---|---|---|---|---|---|---|---|
| CD4+ T cells | CD4+ CD8- | (% of conventional T cells) | 30,623 | 29.2 | 72,970 | 75.7 | 70,332 | 70.2 |
| Treg | CD25+ | (% of CD4+ T cells) | 526 | 1.7 | 3,680 | 5.0 | 4,954 | 7.0 |
| Tfh | CXCR5+ | (% of CD4+ T cells) | 60 | 0.20 | 5,073 | 7.0 | 4,031 | 5.7 |
| **CD4+ naive** | **CD27+ CD45RA+ CD95-** | **(% of CD4+ T cells)** | **3,216** | **10.5** | **33,960** | **46.5** | **19,339** | **27.5** |
| CD4+ Tscm | CD27+ CD45RA+ CD95+ | (% of CD4+ T cells) | 1,066 | 3.5 | 1,222 | 1.7 | 3,890 | 5.5 |
| CD4+ Temra | CD27- CD45RA+ | (% of CD4+ T cells) | 65 | 0.21 | 74 | 0.1 | 72 | 0.1 |
| CD4+ Tem | CD45RA- | (% of CD4+ T cells) | 25,302 | 82.6 | 28,292 | 38.8 | 37,201 | 52.9 |
| CD4+ Tcm | CD45RA- CCR7+ CD28+ | (% of CD4+ T cells) | 17,625 | 57.6 | 22,760 | 31.2 | 32,648 | 46.4 |
| *Marker panel 2* | *singlets live CD45+* | *total cells* | *560,000* | | *560,000* | | *560,000* | |
| B cells | CD19+ CD20+ | (% of total cells) | 37,382 | 6.7 | 74,325 | 13.3 | 36,975 | 6.6 |
| Transitional | CD10+ IgM+ | (% of B cells) | 50 | 0.13 | 1,048 | 1.4 | 1,211 | 3.3 |
| Mature | CD10- | (% of B cells) | 37,332 | 99.9 | 73,277 | 98.6 | 35,764 | 96.7 |
| DN B cells | CD27- IgD- | (% of mature B cells) | 2,650 | 7.1 | 3,072 | 4.2 | 1,386 | 3.9 |
| Naive B cells | CD27- IgD+ | (% of mature B cells) | 33,877 | 90.7 | 58,683 | 80.08 | 26,316 | 73.6 |
| IgD+ CD27+ mature | CD27+ IgD+ | (% of mature B cells) | 352 | 0.94 | 4,303 | 5.9 | 1,169 | 3.3 |
| MZ B cells | CD27+ IgD+ IgM+ | (% of mature B cells) | 93 | 0.25 | 3,134 | 4.3 | 272 | 0.76 |
| $IgD^{only}$ memory | CD27+ IgD+ IgM- | (% of mature B cells) | 225 | 0.60 | 947 | 1.3 | 845 | 2.4 |
| **Memory** | **CD27+ IgD-** | **(% of mature B cells)** | **132** | **0.35** | **6,029** | **8.2** | **6,457** | **18.1** |
| **Plasma cells** | **CD20- IgD- CD27++ CD38++** | **(% of total cells)** | **6** | **0.001** | **102** | **0.018** | **221** | **0.039** |
| pDCs | CD19- CD123+ HLA-DR+ | (% of total cells) | 118 | 0.02 | 1,733 | 0.31 | 2,223 | 0.40 |
| Basophils | CD19- CD123+ HLA-DR- | (% of total cells) | 2,658 | 0.47 | 4,967 | 0.89 | 7,998 | 1.4 |
| Monocytes | CD19- CD123- CD14+ | (% of total cells) | 33,597 | 6.0 | 73,892 | 13.2 | 111,820 | 20.0 |
| classical | CD14+ CD16- | (% of monocytes) | 20,086 | 59.8 | 64,390 | 87.1 | 101,426 | 90.7 |
| **non-classical** | **CD14+ CD16+** | **(% of monocytes)** | **13,511** | **40.2** | **9,502** | **12.9** | **10,394** | **9.3** |
| DCs | CD19- CD123- CD14- HLA-DR+ CD11c+ | (% of total cells) | 14,999 | 2.7 | 4,982 | 0.89 | 6,689 | 1.2 |
| cDC1 | CD141+ CD1c- | (% of total DCs) | 1 | 0.01 | 35 | 0.70 | 37 | 0.55 |
| cDC2 | CD141+ CD1c+ | (% of total DCs) | 1,092 | 7.3 | 4,580 | 91.9 | 5,514 | 82.4 |

ASC, antibody-secreting cells; DC, dendritic cells; DN, double-negative; DP, double-positive; ILCs, innate lymphoid cells; MAIT, mucosal-associated invariant T cells; MZ, marginal zone; PBMCs, peripheral blood mononuclear cells; pDCs, plasmacytoid dendritic cells; Tcm, central memory T cells; Tem, effector memory T cells; Temra, terminally differentiated effector memory T cells; Tfh, follicular helper T cells; Treg, regulatory T cells; Tscm, stem cell-like memory T cells.

**Supplementary Table 3. Single-cell RNA-seq analysis of PBMCs in patient P1 and 7 healthy controls.**

| Cell cluster | Cell type | Patient P1, n | Patient P1, % | all controls, n | all controls, % | 2 adult controls, n | 2 adult controls, % | 5 age-matched controls*, n | 5 age-matched controls*, % |
|---|---|---|---|---|---|---|---|---|---|
| 1-9 | T cells | 4,994 | 60.8 | 25,531 | 58.6 | 10,968 | 58.1 | 14,563 | 59.1 |
| **1** | **Tcm/naive helper T cells** | **198** | **2.4** | **14,010** | **32.2** | **7,632** | **40.4** | **6,378** | **25.9** |
| **2** | **Tcm/naive cytotoxic T cells** | **33** | **0.4** | **5,603** | **12.9** | **1,966** | **10.4** | **3,637** | **14.8** |
| **3** | **Temra cytotoxic T cells** | **1,791** | **21.8** | **1,646** | **3.8** | **35** | **0.2** | **1,611** | **6.5** |
| 4 | Tem/Trm cytotoxic T cells | 111 | 1.4 | 504 | 1.2 | 139 | 0.7 | 365 | 1.5 |
| 5 | Tem/effector helper T cells | 0 | 0 | 521 | 1.2 | 149 | 0.8 | 372 | 1.5 |
| 6 | Regulatory T cells | 403 | 4.9 | 847 | 1.9 | 411 | 2.2 | 436 | 1.8 |
| 7 | Proliferating T cell | 28 | 0.3 | 136 | 0.3 | 23 | 0.1 | 113 | 0.5 |
| **8** | **γδ T cells** | **2,425** | **29.5** | **1,292** | **3.0** | **277** | **1.5** | **1,015** | **4.1** |
| **9** | **MAIT cells** | **5** | **0.1** | **972** | **2.2** | **336** | **1.8** | **636** | **2.6** |
| **10** | **NK cells** | **8** | **0.1** | **224** | **0.5** | **67** | **0.4** | **157** | **0.6** |
| **11** | **CD16+ NK cells** | **26** | **0.3** | **4,289** | **9.9** | **1,358** | **7.2** | **2,931** | **11.9** |
| 12-14 | B cells | 743 | 9.1 | 5,740 | 13.2 | 1,801 | 9.5 | 3,939 | 16.0 |
| 12 | Naive B cells | 612 | 7.5 | 3,896 | 8.9 | 792 | 4.2 | 3,104 | 12.6 |
| 13 | **Memory B cells** | **41** | **0.5** | **1,516** | **3.5** | **952** | **5.0** | **564** | **2.3** |
| 14 | Age-associated B cells | 90 | 1.1 | 328 | 0.8 | 57 | 0.3 | 271 | 1.1 |
| 15 | **Plasma cells** | **0** | **0** | **177** | **0.4** | **102** | **0.5** | **75** | **0.3** |
| 16 | Classical monocytes | 996 | 12.1 | 5,869 | 13.5 | 4,008 | 21.2 | 1,861 | 7.6 |
| 17 | **Non-classical monocytes** | **1,280** | **15.6** | **941** | **2.2** | **177** | **0.9** | **764** | **3.1** |
| 18 | pDC | 2 | 0.02 | 276 | 0.6 | 150 | 0.8 | 126 | 0.5 |
| 19 | DC2 | 9 | 0.1 | 381 | 0.9 | 198 | 1.0 | 183 | 0.7 |
| 20 | HSC/MPP | 49 | 0.6 | 59 | 0.1 | 33 | 0.2 | 26 | 0.1 |
| 21 | Late erythroid | 88 | 1.1 | 27 | 0.1 | 22 | 0.1 | 5 | 0.02 |
| 22 | Megakaryocytes/platelets | 14 | 0.2 | 21 | 0.05 | 9 | 0.05 | 12 | 0.05 |
| 1-22 | Total cells | 8,209 | 100.0 | 43,535 | 100.0 | 18,893 | 100.0 | 24,642 | 100.0 |

* Age-matched controls are subjects NP30, NP31, NP39, NP41 and NP44 from Yoshida et al[8]. PBMCs, peripheral blood mononuclear cells; Tcm, central memory T cells; Tem, effector memory T cells; Temra, terminally differentiated effector memory T cells; MAIT, mucosal-associated invariant T cells; HSC, hematopoietic stem cells; MPP, multipotent progenitors; pDCs, plasmacytoid dendritic cells.
